# Dengue and chikungunya virus transmission in Kinshasa, Democratic Republic of the Congo

**DOI:** 10.64898/2026.06.11.26355423

**Authors:** Rachel Sendor, Jessie Edwards, Kristin Banek, Melchior Mwandagalirwa Kashamuka, Justin Lessler, Daniel O. Espinoza, Izabella N. Castillo, Fabien Vulu, Nono Mvuama, Joseph A. Bala, Marthe Nkalani, Georges Kihuma, Joseph Atibu, Tommy Nseka, Kyaw L. Thwai, Lakshmanane Premkumar, Rhoel R. Dinglasan, Michael Emch, Jonathan J. Juliano, Matthew H. Collins, Antoinette Tshefu, Jonathan B. Parr

## Abstract

Dengue (DENV) and chikungunya (CHIKV) are understudied in the Democratic Republic of the Congo (DRC) and across Africa despite evidence of transmission. We measured DENV and CHIKV IgG seroprevalences in Kinshasa Province, DRC, by antigen-capture ELISA, using dried blood spots from 2021. Force of infection (FOI) was estimated from age-stratified seroprevalences using Bayesian catalytic modeling. Among 1,250 participants, DENV IgG seroprevalence was 38.1% (95% CI: 34.5%-41.8%), increasing with age, and highest within peri-urban Kimpoko sites (54.9%). CHIKV IgG seroprevalence was 24.2% (95% CI: 21.1%-27.6%), increasing with age and comparable between peri-urban Kimpoko and rural Bu, with few seropositives in the city-center. DENV-CHIKV IgG co-occurrence was detected in 12.8% of participants. Time-varying FOI models provided best fit to age-stratified seroprevalences, with spatial variation detected. Sustained DENV and CHIKV circulation across Kinshasa highlights an under-appreciated transmission risk and underscores the need for strengthened arboviral surveillance in the DRC and surrounding region.

## INTRODUCTION

Dengue virus (DENV) and chikungunya virus (CHIKV) are primarily transmitted by *Aedes aegypti* and *Ae. albopictus* mosquito vectors and pose emerging global health threats. DENV is the most prevalent arboviral infection globally and co-circulates with CHIKV across tropical and subtropical regions, with models estimating billions of people reside in areas at risk of transmission(*1–3*). Most infections are asymptomatic or cause mild, self-limiting febrile illness; overall mortality is low(*4,5*). However, severe dengue develops in approximately 5% of infections and carries high risk of hospitalization and morbidity(*4*). CHIKV elicits prolonged, debilitating joint pain in a substantial proportion of cases and severe illness can occur, particularly in high-risk groups including neonates, elderly, and those with comorbidities(5–7). Large-scale outbreaks of both viruses strain health systems and cause economic hardship in affected communities(8–13). DENV and CHIKV circulate widely in Africa, yet epidemiological characterization remains limited. Dengue is endemic across ≥30 African countries, with circulation of DENV 1-3 serotypes and reports of DENV-4(14–17). CHIKV is similarly widespread, with documented regional transmission and recurrent outbreaks(18). Available evidence largely derives from small serosurveys, outbreak investigations, or returning travelers. As a result, fundamental aspects of DENV and CHIKV epidemiology in Africa, including transmission intensity, spatial heterogeneity, and population-level immunity, remain largely unknown.

The Democratic Republic of the Congo (DRC) is the largest country in sub-Saharan Africa, and its capital, Kinshasa, is one of the continent’s largest cities. DENV was first reported in the DRC in 1960, and CHIKV in 1958, with CHIKV outbreaks reported in Kinshasa in 1999, 2012, and 2019(19). DENV seropositivity has been identified through limited studies among Kinshasa residents and returning travelers(19–25). *Ae. aegypti* and *Ae. albopictus* are well-established in the region. DENV has been isolated from mosquito pools, indicating active transmission potential(26–29). However, absence of sustained national surveillance for these viruses, limited diagnostic capacity, and overlapping clinical presentations with malaria and other prevalent febrile illnesses obscure accurate detection and estimation of disease burden in the population.

We aimed to characterize DENV and CHIKV transmission in Kinshasa Province using samples collected across three health areas, measuring seroprevalence, identifying correlates of prior infection, and estimating force of infection (FOI).

## METHODS

### Study design

This cross-sectional serological study used survey data and residual dried blood spots (DBS) from the final household visit of a longitudinal malaria cohort study in Kinshasa Province(30). The parent cohort (phase II) began in April 2018, with the endline survey during the 2021 rainy season. The final survey population comprised children ≥3 years of age and adults from seven sites across three health areas. Loss to follow-up between the parent cohort and analytic sample was 23% **Supplemental Figure 1**.

Site and household sampling has been previously described(30). Briefly, sites were selected using stratified sampling by health area to capture variation in environment, malaria endemicity, and urbanicity, including three sites (Bu, Impuru, and Pema) in rural, arid, grassy highlands, three sites (Kimpoko, Ngamanzo, and Iye) within peri-urban, low-lying marshlands along the Congo River, and one highly-urban site, Voix du Peuple, in the city center (**Figure 1**)(30). Individual-level and household-level surveys were conducted alongside DBS collection (**Supplemental Table 1)**.

**Figure 1.**
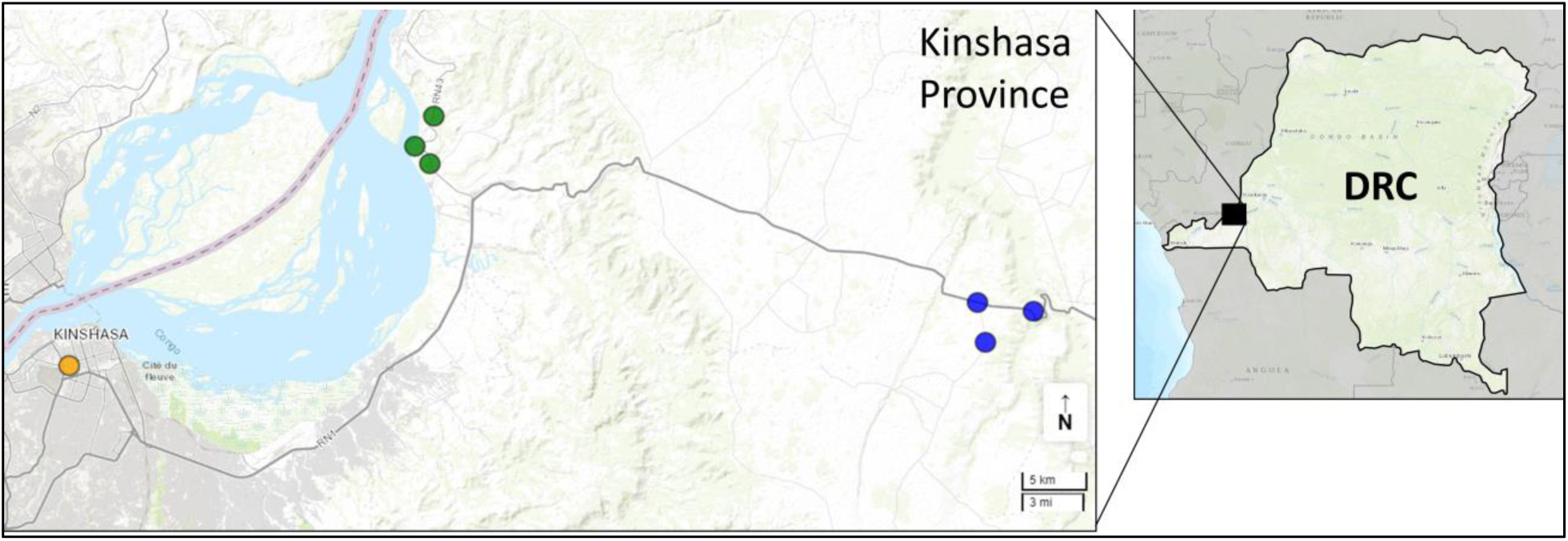
Study sites across an urban-rural gradient within Kinshasa Province, DRC. Participants were enrolled by household across seven sites selected for the study: one urban site in Voix du Peuple in downtown Kinshasa (orange), three peri-urban sites in the Kimpoko health area located along the Congo River (green), and three rural sites in the Bu health area located in the grassy highlands (blue). Roads are depicted in gray. Source: *ESRI*(31).

### Antigen capture ELISA (primary endpoint)

We conducted high-throughput, semi-automated antigen capture enzyme-linked immunosorbent assay (ELISA) on DBS to assess serostatus(32). Plasma proteins were eluted in 2022 from three 6mm punches per DBS, and eluant was stored at −80°C. Plates were coated with capture antibodies (flavivirus cross-reactive monoclonal antibody [4G2] for DENV, alphavirus-binding mouse monoclonal antibody [CHK-48] for CHIKV) overnight, washed with Tween-PBS and blocked with 3% milk. For DENV, antigen comprised serotypes 1-4 derived from cleared C6/36 culture supernatants; for CHIKV, antigen was supernatant from Vero cell cultures infected with attenuated strain 181/clone25(33,34). Plates were incubated, washed, and eluants added in duplicate. Alkaline phosphatase (AP)-conjugated goat anti-human IgG (Sigma Aldrich, Co. Cat# A9544) was applied, and plates were developed with p-nitrophenylphosphate (pNPP) substrate (Sigma Aldrich, Co. Cat# N1891-50SET). Optical density (OD; 405nm) was measured every 5 minutes for 30 minutes. IgG-seropositivity cut-offs were defined by background subtraction per-plate, excluding lowest OD and using mean of remaining 5 lowest samples + 3 standard deviations + 0.3 inflation-adjustment factor(22).

### Orthogonal DENV serological testing

A 20% age-stratified random subset of DBS underwent orthogonal DENV testing in a separate laboratory using: 1) ELISA using recombinant, covalently-cross-linked, stabilized DENV-2 envelope protein (E) dimers(35) lacking pre-membrane protein (prM) and with reduced exposure of the E protein fusion loop, and 2) multiplex bead-based assay (BBA) using E domain III (EDIII) antigens from DENV 1-4(36). Younger ages were over-sampled (50% of 3-6 and 7-10 year-olds and 10% of ≥11 year-olds were randomly sampled) to enhance resolution of recent transmission.

### DENV anti-E dimer ELISA

Plates were coated with recombinant E-dimer antigen (4 µg/ml in 1xTBS), blocked with 3% milk and tested for human anti-dengue IgG binding with freshly-extracted serum from DBS. pNPP substrate hydrolysis by AP-conjugated detection antibody was measured at 405 nm (OD) and a 0.2 OD assay threshold was applied using naïve specimens. Agreement between E-dimer and whole virus antigen capture ELISAs was measured using Cohen’s kappa.

### EDIII BBA

Samples in the orthogonal testing subset seropositive by E-dimer ELISA were further tested by EDIII BBA. Site-specifically biotinylated DENV1-4 and Halo-tag background control were coupled to MagPlex-Avidin microspheres(36). Antigen-coupled beads were incubated with 1:500 human serum from DBS, then washed and incubated with PE-conjugated goat anti-human IgG Fc secondary antibody, and fluorescence analysis performed using Luminex 200^®^. Samples with high background noise to Halo-tag-coupled beads (>1000) were excluded. Without reference specimens from African settings, previously established thresholds from non-African settings were applied (EDIII: DENV1 (>1306 MFI), DENV2 (>1082 MFI), DENV3 (>1364 MFI), and DENV4 (>1285 MFI)) to inform seropositivity(36); lack of African-specific thresholds and uncertainty in background flaviviruses and antigenic diversity among strains in the region precluded reliable serotyping.

### Statistical Analysis

Primary outcomes in this study were DENV IgG and CHIKV IgG seroprevalences, estimated overall, and by age, health area, and site. Inverse probability weights addressed loss-to-follow-up from the parent cohort baseline, conditional on age, site, and wealth. Robust 95% confidence intervals (CIs) accounted for household-level clustering and weighting. Associations between DENV and CHIKV seropositivity were assessed by chi-square, or fisher’s exact for small sample sizes. Seropositive results reflect probable infection due to potential for cross-reactivity.

We estimated prevalence ratios for correlates of infection history using unadjusted modified Poisson regression. Each factor was modeled separately. Age groups were more granular for seroprevalence estimation to capture detail and recent transmission, and aggregated for prevalence ratio modeling due to sample size.

We estimated population-level transmission intensity using catalytic FOI modeling, where FOI, (λ), represents the hazard rate of infection among susceptibles, derived from age-specific seroprevalences(37). Constant transmission and time-varying models were compared using leave-one-out cross-validation information criteria (LOOIC). Constant models assume no variation in transmission intensity across time or age. Time-varying models allow variation over time, though assume constant FOI within 5-year intervals and age. Both models assume people are susceptible and seronegative at birth and no sero-reversion; DENV estimates represent hazard rate of first infection. Bayesian Markov Chain Monte Carlo models were run using 4 chains and 2,000 iterations (500 warm-up) per model. Convergence was assessed using R-hat. Median FOI and 95% credible intervals (CrI) were sampled from posterior distributions using weakly-informative priors. We investigated divergent chains in time-varying stratified models due to small sample sizes, and stabilized sampling with strengthened smoothing parameters and priors informed from total population. Ages ≥71 were collapsed to a single category due to sparse samples. See **Supplemental Table 2** for details.

Sensitivity analyses quantified potential misclassification from ELISA cross-reactivity with non-DENV flaviviruses, and non-CHIKV alphaviruses. For DENV, analyses were repeated in the orthogonally-tested subset, among samples seropositive by both whole-virus and recombinant E-dimer ELISAs; age-standardization accounted for stratified sampling. DENV IgG seropositivity across all three assays was also described. Seroprevalences and FOI models for DENV and CHIKV were also re-analyzed under 20%-40% false positivity scenarios(38), reclassifying a random subset of seropositives as IgG-seronegative, assuming non-differential outcome misclassification.

Missing data were minimal and imputed by multiple imputation where applicable, using all available data to inform imputations (**Supplemental Table 3**). Those with missing outcome data were excluded from FOI and sensitivity analyses.

Analyses were conducted in R (version 4.2). All R packages used in this analysis are included in **Supplemental Table 4.**

### Ethical Review

Informed consent, and assent where required, were obtained prior to enrollment and sampling. The parent cohort and present study were approved by Institutional Review Boards (IRB) at the University of North Carolina at Chapel Hill (IRB#’s: 17-1588; 25-0096) and Kinshasa School of Public Health (IRB#’s: ESP/CE/021/2017; ESP/CE/328/2025).

## RESULTS

### Study population

A total of 1,250 participants with biospecimens across 219 households were included, representing 76% of baseline participants (92% of households). Among these, 1,138 participants provided survey data at sampling. Participants ranged in age from 3-87 years; 38% (n=475) lived within rural Bu, 39% (n=488) within peri-urban Kimpoko, and 23% (n=287) in downtown, urban Voix du Peuple. Forty-seven percent had a secondary/post-secondary education, 28% lived in households with structural openings/holes, 22% reported history of recent travel, and 20% lived near water (**Table 1**). Participant demographics by health area are shown in **Supplemental Table 3**.

**Table 1.** Participant characteristics.

| Participant characteristics | Study sample<br>(unweighted) | Weighted to target<br>population* |
| --- | --- | --- |
| No. (%) | 1,250 (100) | (100) |
| Health area |  |  |
| Bu (rural) | 475 (38.0%) | (34.8%) |
| Kimpoko (peri-urban) | 488 (39.0%) | (39.2%) |
| Voix du Peuple (urban) | 287 (23.0%) | (26.0%) |
| Village site |  |  |
| Bu | 163 (13.0%) | (11.7%) |
| Impuru | 166 (13.3%) | (12.2%) |
| Pema | 146 (11.7%) | (10.9%) |
| Ngamanzo | 201 (16.1%) | (15.7%) |
| Iye | 107 (8.6%) | (8.5%) |
| Kimpoko | 180 (14.4%) | (15.0%) |
| Voix du peuple | 287 (23.0%) | (26.0%) |
| Sex |  |  |
| Female | 676 (54.1%) | (54.1%) |
| Age (years) |  |  |
| Median (IQR) | 18 (10, 38) | (10, 37) |
| Range | 3 - 87 | 3 - 87 |
| Education attained |  |  |
| No school | 104 (8.3%) | (8.1%) |
| Primary | 473 (37.8%) | (37.1%) |
| Secondary / Post-secondary | 581 (46.5%) | (47.7%) |
| Other | 33 (2.6%) | (2.6%) |
| Under school-age (<5 years) | 59 (4.7%) | (4.6%) |
| SES (wealth) score |  |  |
| Poorest | 237 (19.0%) | (17.4%) |
| Poorer | 240 (19.2%) | (17.9%) |
| Average wealth | 256 (20.5%) | (20.0%) |
| Wealthier | 271 (21.7%) | (22.4%) |
| Wealthiest | 246 (19.7%) | (22.4%) |
| Num. household members |  |  |
| Median (IQR) | 7 (5, 9) | (5, 9) |
| Mosquito nets in household |  |  |
| Yes | 981 (78.5%) | (78.3%) |
| Num. mosquito nets in household |  |  |
| Median (IQR) | 2 (1, 3) | (1, 3) |
| Water w/in 2 min. walk |  |  |
| Yes | 243 (19.4%) | (19.5%) |
| Type of water source |  |  |
| Stream | 58 (4.6%) | (4.8%) |
| Pond/lake | 28 (2.3%) | (2.1%) |
| Swamp | 13 (1.1%) | (1.0%) |
| Frequent puddles | 113 (9.0%) | (8.9%) |
| Other | 31 (2.5%) | (2.7%) |
| No water source | 1007 (80.6%) | (80.5%) |
| Open eaves/holes in home |  |  |
| Yes | 347 (27.8%) | (26.4%) |
| Owns cultivable land |  |  |
| Yes | 345 (27.6%) | (26.0%) |
| Window material |  |  |
| Glass | 196 (15.7%) | (17.6%) |
| Screens | 9 (0.7%) | (0.7%) |
| Plastic | 38 (3.0%) | (2.8%) |
| Planks | 480 (38.4%) | (37.9%) |
| Open holes | 152 (12.2%) | (11.3%) |
| Other | 264 (21.1%) | (21.5%) |
| No windows | 3 (0.2) | (0.2%) |
| History of travel |  |  |
| Yes | 279 (22.3%) | (21.6%) |

\*Weighting addresses selection into the present study population to represent characteristics of the parent cohort study in order to better reflect Kinshasa Province.

### Seroprevalence

Overall DENV IgG seroprevalence was 38.1% (95% CI: 34.5%–41.8%), CHIKV IgG seroprevalence was 24.2% (95% CI: 21.1%-27.6%), and DENV-CHIKV co-seropositivity was 12.8% (95% CI: 10.9%-15.1%) (**Table 2**). DENV seroprevalence increased with age, from 14.9% among 3-5 year olds, to 68.5% among adults >50. CHIKV seroprevalence fluctuated across ages, lowest among children aged 3-5 (8.1%), similar between ages 6-15 (14.6%) and 16-25 (14.2%), and highest among adults >50 (53.5%).

**Table 2.** DENV and CHIKV IgG seroprevalence in Kinshasa Province, DRC*^,†^.

|  | DENV (IgG) seroprevalence |  |  | CHIKV (IgG) seroprevalence |  |  | DENV & CHIKV co-seroprevalence |  |  |
| --- | --- | --- | --- | --- | --- | --- | --- | --- | --- |
|  | n | % | 95% CI | n | % | 95% CI | n | % | 95% CI |
| Total | 484 | 38.1 | 34.5 – 41.8 | 327 | 24.2 | 21.1 – 27.6 | 173 | 12.8 | 10.9 – 15.1 |
| Age (years) |  |  |  |  |  |  |  |  |  |
| 3 - 5 | 14 | 14.9 | 8.0 – 21.8 | 10 | 8.1 | 2.5 – 13.7 | 2 | 2.1 | -1.6 – 5.8 |
| 6 - 15 | 110 | 25.3 | 20.5 – 30.0 | 79 | 14.6 | 10.2 – 18.9 | 22 | 4.6 | 2.4 – 6.8 |
| 16 - 25 | 95 | 38.9 | 32.7 – 45.2 | 31 | 14.2 | 10.0 – 18.4 | 16 | 6.6 | 3.5 – 9.7 |
| 26 - 50 | 154 | 54.0 | 47.8 – 60.1 | 122 | 43.0 | 36.4 – 49.5 | 73 | 25.4 | 19.9 – 31.0 |
| 51+ | 111 | 68.5 | 60.9 – 76.1 | 86 | 53.5 | 44.8 – 62.1 | 60 | 37.2 | 29.2 – 45.2 |
| Health area |  |  |  |  |  |  |  |  |  |
| Bu (rural) | 120 | 24.3 | 20.3 – 28.4 | 156 | 31.3 | 25.5 – 37.1 | 52 | 10.2 | 7.4 – 13.1 |
| Kimpoko (peri-urban) | 275 | 54.9 | 49.4 – 60.3 | 157 | 29.9 | 25.3 – 34.5 | 115 | 21.9 | 18.4 – 25.3 |
| Voix du Peuple (urban) | 89 | 29.7 | 23.0 – 36.4 | 14 | 4.6 | 2.1 – 7.1 | 6 | 1.9 | 0.4 – 3.4 |
| Study site |  |  |  |  |  |  |  |  |  |
| *Bu* |  |  |  |  |  |  |  |  |  |
| Bu | 41 | 24.4 | 17.8 – 31.0 | 38 | 21.5 | 13.8 – 29.3 | 13 | 7.2 | 3.9 – 10.5 |
| Impuru | 47 | 27.3 | 19.4 – 35.1 | 39 | 21.7 | 15.2 – 28.3 | 17 | 9.2 | 5.4 – 13.1 |
| Pema | 32 | 21.0 | 14.7 – 27.3 | 79 | 53.2 | 43.8 – 62.6 | 22 | 14.6 | 7.9 – 21.3 |
| *Kimpoko* |  |  |  |  |  |  |  |  |  |
| Ngamanzo | 124 | 60.6 | 52.3 – 68.8 | 58 | 26.5 | 20.7 – 32.2 | 47 | 21.6 | 16.3 – 26.9 |
| Iye | 41 | 36.0 | 26.5 – 45.4 | 41 | 37.0 | 23.6 – 50.4 | 23 | 20 | 12.6 – 27.4 |
| Kimpoko | 110 | 59.9 | 52.0 – 67.8 | 57 | 29.8 | 23.4 – 36.1 | 45 | 23.4 | 17.7 – 29.1 |
| *Voix du Peuple* | 89 | 29.7 | 23.0 – 36.3 | 14 | 4.6 | 2.1 – 7.1 | 6 | 1.9 | 0.4 – 3.4 |

\* Seroprevalence was estimated in the weighted population to account for nonrandom
selection into the study sample.
† IgG+ sample counts include 10 imputed DENV and CHIKV outcomes.

Seroprevalence also varied by site. DENV seroprevalence was highest in peri-urban Kimpoko (54.9% [95% CI: 49.4%–60.3%%]), followed by urban Voix du Peuple (29.7% [95% CI: 23.0%–36.4%]), and rural Bu (24.3% [95% CI: 20.3%–28.4%]) (**Table 2**). CHIKV seropositivity was similar in Kimpoko (29.9% [95% CI: 25.3%–34.5%]) and Bu (31.3% [95% CI: 25.5%–37.1%]), and lowest in Voix du Peuple (4.6%; 95% CI: 2.1%–7.1%).

DENV-CHIKV co-seropositivity was most common in Kimpoko (21.9% [95% CI: 18.4%-25.3%]), and infrequent within urban Voix du Peuple (1.9% [95% CI: 0.4%-3.4%]). DENV – CHIKV co-seropositivity overall was greater than would be expected by chance (χ2 = 37.56; p<0.001). This association held among peri-urban Kimpoko (χ2 =26.87; p<0.001) and rural Bu sites (χ2 = 8.01; p<0.01), but not within urban Voix du Peuple (p=0.24).

### Force of infection

For DENV, we estimated a 0.022 (95% CrI: 0.020–0.025) annualized per-capita rate of first DENV infection, assuming constant transmission (**Table 3, Supplemental Figure 5**). DENV FOI varied by health area, ranging from 0.014 (95% CrI: 0.012–0.017) in rural Bu, 0.015 (95% CrI: 0.012-0.019) in urban Voix du Peuple, and 0.041 (95% CrI: 0.036–0.046) in peri-urban Kimpoko. Time-varying models fit to age-stratified seroprevalence curves indicate DENV FOI increased during recent intervals, reaching an estimated FOI of 0.032 during the 2016-2021 period (**Figures 2 and 3**); this model-estimated increase was driven primarily by Kimpoko (2016-2021: 0.053) (**Supplemental Figure 6)**.

**Figure 2.**
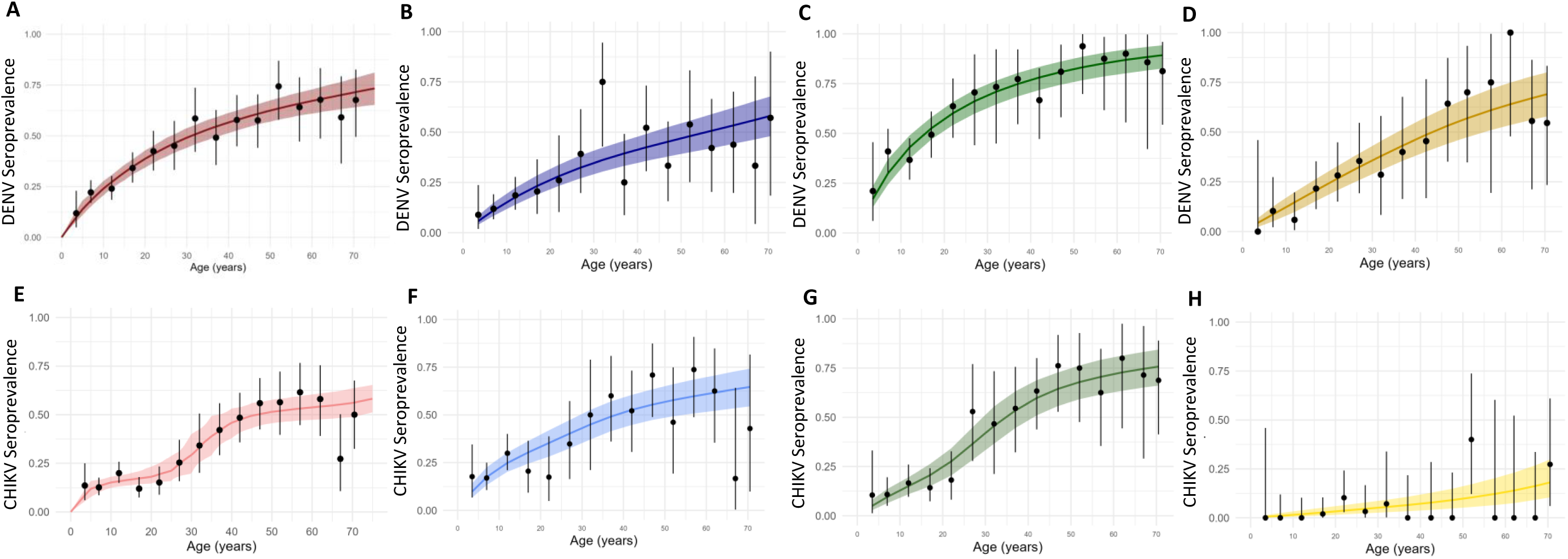
DENV (a-d) and CHIKV (e-h) age-stratified seroprevalences, modeled overall and by site health area, assuming varying transmission intensity by age and calendar time. DENV (a-d) and CHIKV (e-h) age-stratified seroprevalences within 10-year age strata are plotted on the primary y-axis for observed (*points and 95% CIs*) and posterior-predicted (*solid line and 95% CrIs*) values for time-varying FOI models. Age-stratified seroprevalence curves are presented for the total population (**red**), and by Bu health area sites (**blue**), Kimpoko health area sites (**green**), and downtown Kinshasa site (**yellow**). Time-varying transmission models used birth year and year of survey to capture time-at-risk within a catalytic modeling function, allowing fluctuation over time, and assume no waning immunity among infected individuals. Samples sizes and seropositive sample counts varied across site and age strata, as presented in **Table 2**. FOI: force of infection; CI: confidence interval; CrI: credible interval.

**Figure 3.**
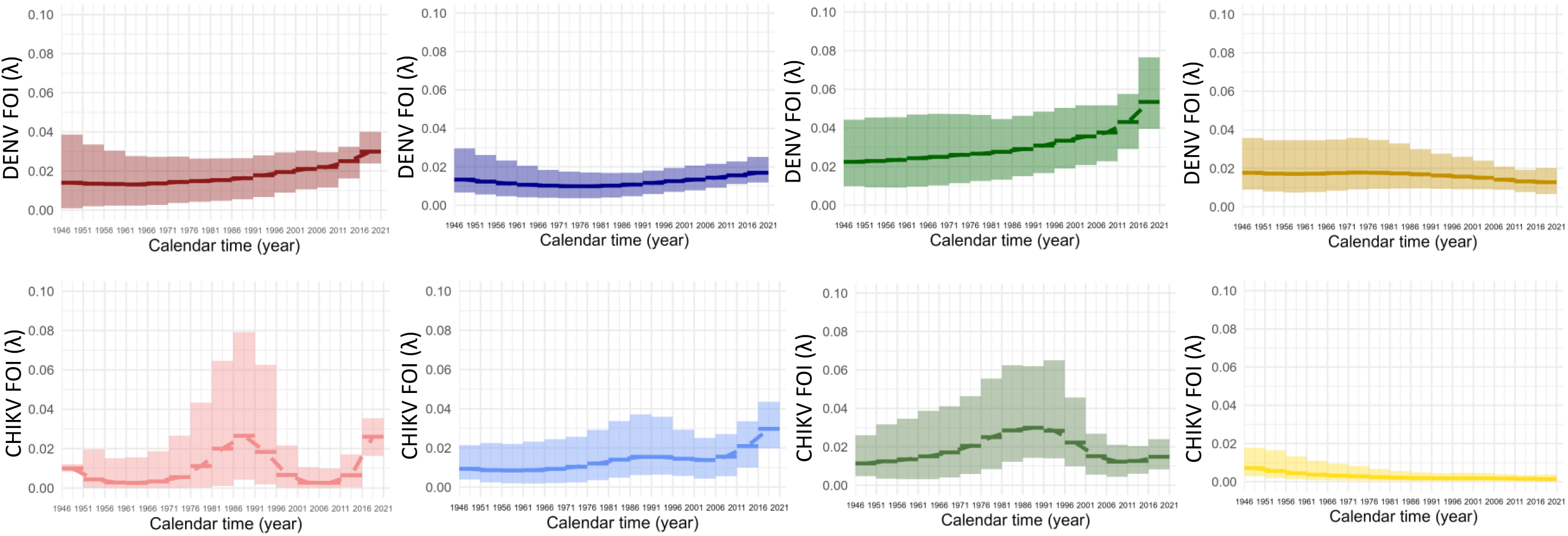
DENV and CHIKV force of infection, modeled overall and by site health area, assuming varying transmission intensity by age and calendar time. DENV (a-d) and CHIKV (e-h) force of infection (FOI), predicted from posterior distributions (*dashed line and colored 95% CrIs*) are plotted on the y-axis for the total population (**red),** and by Bu health area sites (**blue**), Kimpoko health area sites (**green**), and downtown Kinshasa site (**yellow**). Time-varying transmission models used birth year and year of survey to capture time-at-risk within a catalytic modeling function, allowing FOI to fluctuate by 5-year piecewise intervals, and assume no waning immunity among infected individuals. Corresponding FOI estimates and comparisons for model fit are presented in **Table 3**. Samples sizes and seropositive sample counts varied across site and age strata, as presented in **Table 2**. FOI: force of infection; CI: confidence interval; CrI: credible interval

**Table 3:**
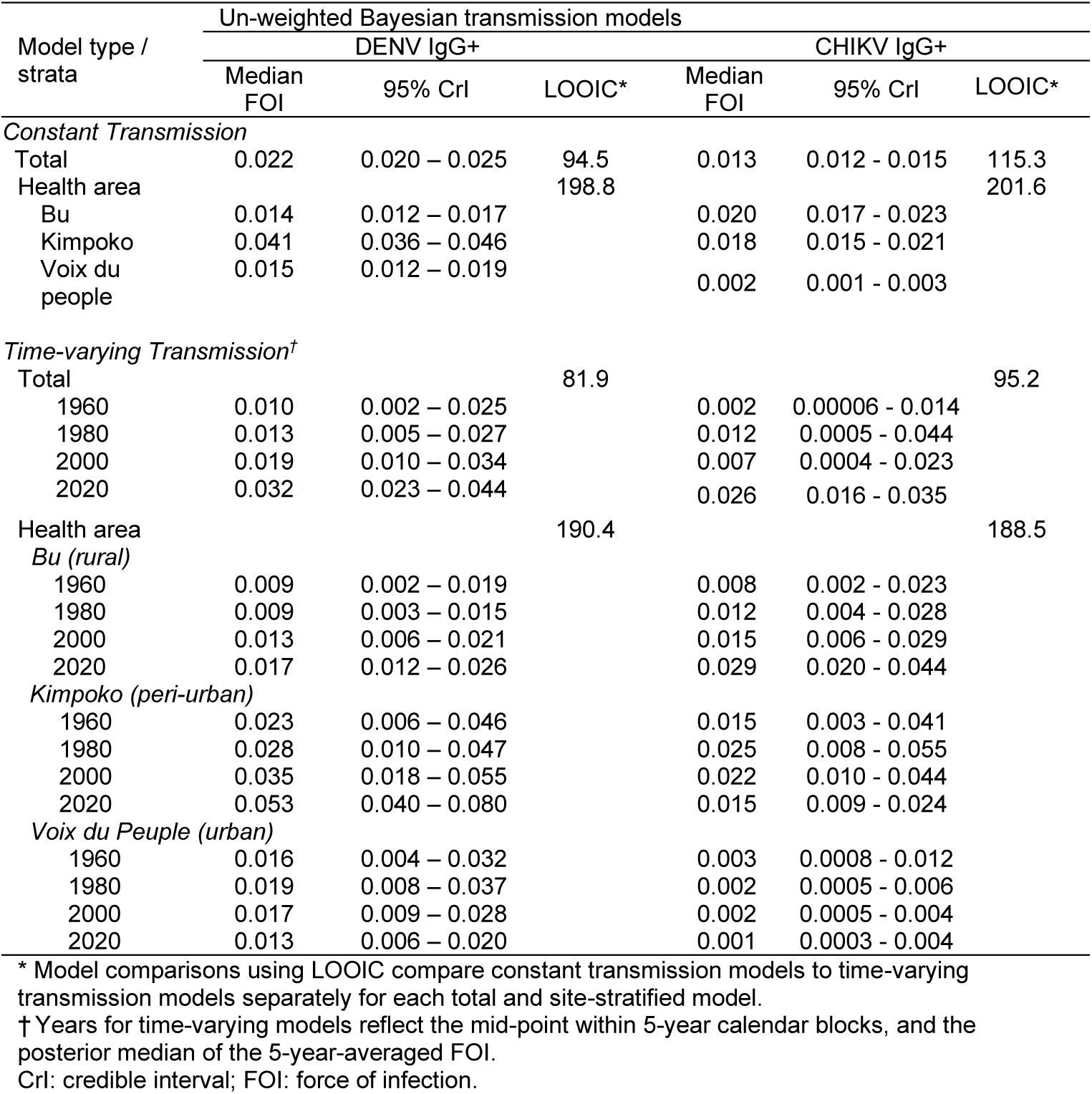
DENV & CHIKV force of infection transmission models.

For CHIKV, we estimated a 0.013 (95% CrI: 0.012-0.015]) annualized per-capita rate of infection, assuming constant transmission (**Table 3, Supplemental Figure 5**). CHIKV FOI was similar between Bu and Kimpoko (0.020 [95% CrI: 0.017-0.023] vs. 0.018 [95% CrI: 0.015-0.021]), and lowest in urban Voix du Peuple (0.002 [95% CrI: 0.001-0.003). Time-varying models fit to age-stratified CHIKV seroprevalences estimated two peaks in FOI, the first driven by Kimpoko, and the second by Bu within recent intervals (**Figures 2 and 3**). FOI within Voix du Peuple declined slightly over time, though the low number of CHIKV seropositive samples in this site (n=14) limits interpretability. Time-varying models provided best fit to age-stratified seroprevalences for both DENV and CHIKV, based on lower LOOICs.

### Correlates of infection history

DENV and CHIKV seroprevalences were similar between men and women (1.08 [95% CI: 0.95-1.24] and 1.10 [95% CI: 0.91-1.33], respectively) (**Figure 4**). Increasing age was associated with higher DENV and CHIKV seroprevalences: adults >50 years had 3.22 (95% CI: 2.42–4.29) times the DENV seroprevalence and 5.12 (95% CI: 3.32–7.91) times the CHIKV seroprevalence of that in children 3-9 years. Health area was associated with seropositivity, with distinct viral-specific patterns. DENV seroprevalence in Kimpoko was 2.25 (95% CI: 1.85–2.74) times the seroprevalence in rural Bu, though less pronounced in Voix du Peuple vs. Bu (PR = 1.22 [95% CI:0.92-1.62]). Alternatively, CHIKV seroprevalence was similar in Kimpoko and Bu (0.96 [95% CI: 0.75– 1.22]), and lower in Voix du Peuple (PR: 0.15 [95% CI: 0.08-0.26]).

**Figure 4.**
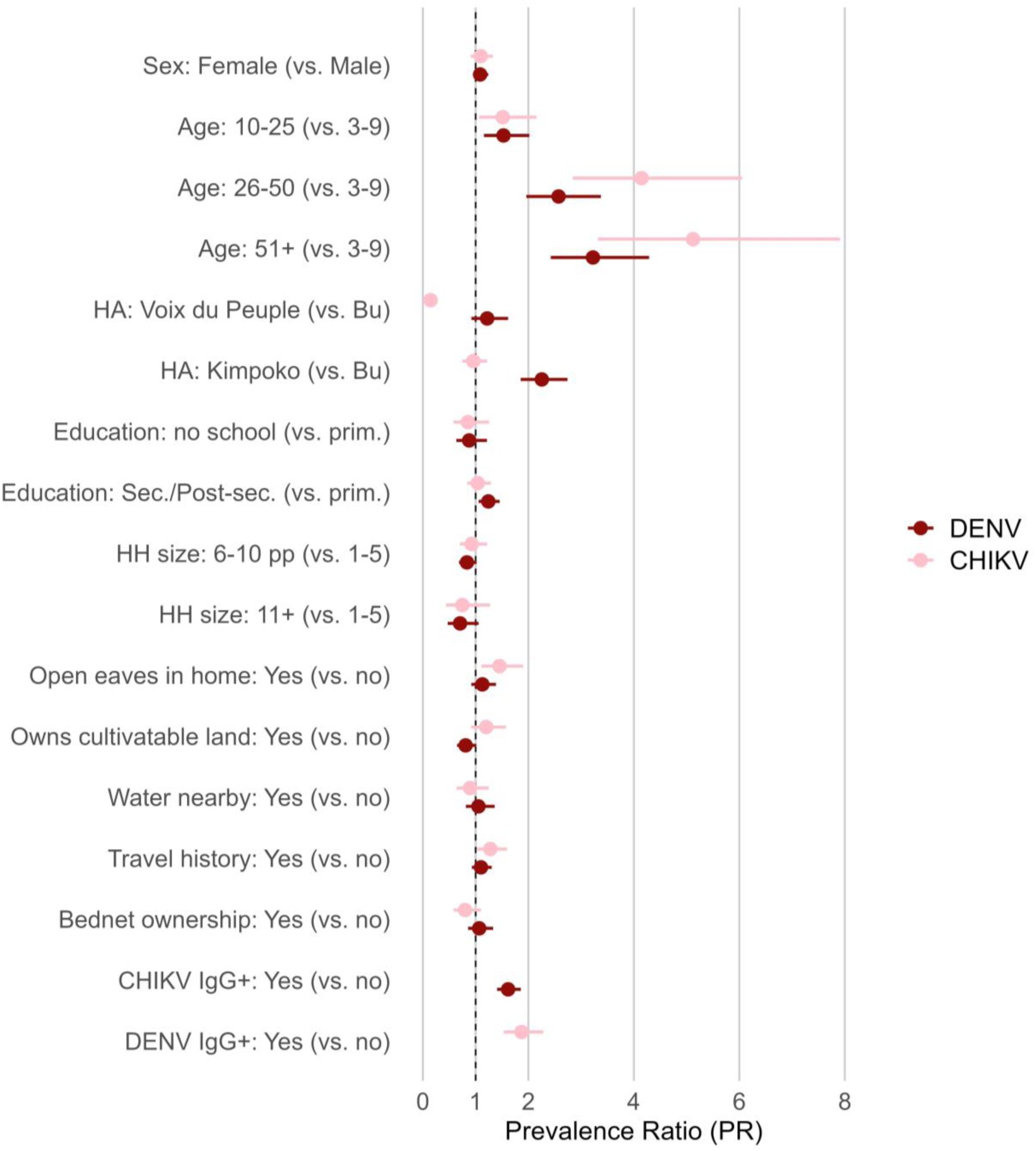
Correlates of DENV and CHIKV IgG seropositivity. Factors associated with DENV IgG and CHIKV IgG seropositivity by whole virus ELISA. Children <5 years ineligible to attend school are excluded from education factors. PRs <1.0 are protective. HA: health area; Prim: primary education; Sec../ Post sec.: secondary / post-secondary education.

Participants with no schooling had lower DENV seroprevalence (0.88 [95% CI: 0.63-1.21]) and CHIKV seroprevalence (0.85 [95% CI 0.58-1.26]) than those with primary-level education, whereas participants with secondary/post-secondary-level educations had higher DENV seroprevalences (1.24 [95% CI: 1.06-1.46]), yet similar CHIKV seroprevalences (1.04 [95% CI: 0.84-1.29]) than those with primary education.

Those living in houses with open eaves/holes had 1.13 (95% CI: 0.92-1.39) times the DENV seroprevalence, and 1.45 (95% CI: 1.11-1.90) times the CHIKV seroprevalence compared to those living in houses without. Participants reporting recent travel outside the village had similar DENV (1.10 [95% CI: 0.93-1.31]) and higher CHIKV (1.28 [95% CI: 1.02-1.60]) seroprevalences to those without travel.

DENV and CHIKV seropositivity were associated. CHIKV IgG seropositive participants had 1.62 (95% CI: 1.41-1.86) times the DENV seroprevalence vs. CHIKV seronegatives, and correspondingly, DENV IgG seropositive participants had 1.87 (95% CI: 1.53-2.28) times the CHIKV seroprevalence vs. DENV IgG seronegatives.

Prevalence ratios stratified by health area are in **Supplemental Figures 3 and 4**.

### DENV orthogonal testing

A 260-sample subset was selected for DENV IgG cross-evaluation. Among 83/260 samples DENV IgG seropositive by whole-virus ELISA, 66% (n=55) were also E-dimer ELISA IgG seropositive, with high concordance between assays (Cohen’s ĸ = 0.69) (**Supplemental Table 5**). Restricting to samples seropositive by both ELISAs yielded an age-standardized seroprevalence of 30.2% (95% CI: 22.7%–37.6%; n=55/260), maintaining age and geographic patterns observed in the primary analysis, with highest seroprevalence still in peri-urban Kimpoko (37.2%, 95% CI: 26.4%–47.9%; n=38), followed by Voix du Peuple (19.1%, 95% CI: 6.1%–32.3%; n=7) and Bu (8.0%, 95% CI: 2.7%–13.2%; n=10) (**Supplemental Table 6**).

Subsequent testing of E-dimer ELISA seropositives by EDIII BBA yielded 30/58 (52%) samples that met positivity thresholds; 28/30 were also whole-virus ELISA seropositive (**Supplemental Figure 2**). The remaining 28/58 (48%) samples were below EDIII thresholds. Since Africa- and DRC-specific thresholds are not available, we cannot exclude the possibility that non-DENV flavivirus antibodies in this population bind to DENV antigens under the dilution tested, warranting further evaluation.

### Sensitivity analysis

DENV IgG seroprevalence decreases to 30.2% - 22.8% under 20% - 40% false positivity scenarios, with proportional declines across sites. FOI estimates similarly decreased (0.016-0.011) assuming 20%-40% false positivity, with overall temporal and spatial patterns unchanged from primary analysis (**Supplemental Tables 7 & 8)**. CHIKV IgG seroprevalence declined to 19.6%-14.7%, respectively, assuming CHIKV misclassification scenarios of 20%-40%, with corresponding reductions in FOI (0.01-0.007 overall), and no change in primary analysis trends (**Supplemental Tables 9 & 10).** Full sensitivity analysis results are presented in **Supplemental Tables 8 & 10**.

## DISCUSSION

We demonstrate historical and recent transmission of DENV and CHIKV in Kinshasa Province, with distinct transmission patterns across sites. Past arboviral infection was common, with 38% of participants DENV IgG seropositive, and 24% CHIKV IgG seropositive, including 13% DENV and CHIKV seropositive. Peri-urban Kimpoko sites within low-lying marshlands along the Congo River had highest DENV seroprevalences, with estimated DENV FOI nearly three times higher than other sites, suggesting localized environmental, behavioral and/or vector-related conditions could be contributing to increased transmission.

CHIKV patterns differed from DENV, with low seropositivity and FOI within highly-urban Voix du Peuple, but higher and comparable seroprevalences in rural Bu and peri-urban Kimpoko. Low CHIKV seroprevalence in Voix du Peuple may reflect the higher household wealth, higher education-level, older population, and lower reported travel characteristics of this site; however, infrequent infections constrained identification of site-specific correlates. Notably, these factors may not similarly impact DENV risk within this same urban population. The higher DENV seroprevalence in this site suggests virus-specific transmission-related factors across sites, with potential implications for risk stratification and prevention.

Spatial heterogeneity in DENV and CHIKV transmission may be driven in part by shifting *Aedes* ecology. *Ae*. *albopictus* has become increasingly dominant in the DRC, favoring peri-urban settings with increased vegetation over urban environments(21,26). Contemporaneous surveillance at these sites detected primarily *Ae. aegypti* in highly urban Kinshasa sites, but little to no *Ae. albopictus*, despite detecting widespread *Ae. albopictus* elsewhere(26). Differences in *Aedes* species-specific vector competence could also contribute. Notably, CHIKV strains detected in the 2019 Kinshasa outbreak harbored the A226V mutation, which increases viral transmissibility by *Ae. albopictus*(26,39).

DENV seroprevalence in our study exceeds estimates from prior investigations in the DRC. A 2013 national survey of children <5 years old reported low seropositivity (0.4% by neutralizing antibody assay; 3.8% by ELISA), pooled across regions(22), compared with 15% among 3-5 year olds in our study. Smaller, Kinshasa-based studies among febrile patients reported DENV seroprevalence near 30% by ELISA, comparable to 38% seroprevalence observed here(20,21). Our larger and age-diverse sample, inclusion of non-febrile participants, and sampling beyond the downtown urban setting, differentiates our study and likely accounts for the higher DENV seroprevalences observed. CHIKV IgG seropositivity was consistent with estimates from prior CHIKV serological testing in Kinshasa (26% among febrile patients in 2015-16)(20). However, our sampling period captures increased population-level immunity following the documented 2019 CHIKV outbreak(39).

Time-varying FOI estimates suggest historic and ongoing low-level endemic DENV transmission in urban Voix du Peuple, while patterns consistent with increasing transmission were observed in peri-urban Kimpoko during the most recent modeled intervals. Despite this increase, annualized risk of DENV infection remained relatively low overall (∼1%-5%). CHIKV displayed a distinct transmission profile, with two suggestive peaks over time alongside ongoing low-level transmission in select sites; transmission in Voix du Peuple remained consistently low or absent. Prior CHIKV outbreaks were reported in Kinshasa in 1999-2000, 2012, and 2019(39–41). While our models do not capture fine-scale annual fluctuations, and were not designed to reconstruct specific outbreak dynamics, they do reflect increased population-level CHIKV immunity among Kimpoko participants after 1995-2000, and a higher model-estimated FOI during 2016-2021 temporally consistent with the 2019 outbreak. Few studies have quantified arboviral FOI in African settings, though a global review applied catalytic models to published seroprevalence data and reported comparable DENV FOI estimates (ranging from 0.004 [Kenya] to 0.11 [Tanzania])(42). Higher 20% annual per-capita infection risk was reported during a 2016 DENV outbreak in Burkina Faso(43). Similar catalytic modeling for CHIKV across African populations demonstrates both endemic and sporadic, epidemic transmission, with geographic variation(44). Detected DENV IgG seropositivity in our study among children <9 years, and overall low FOI, do not meet current WHO-recommended high-transmission thresholds for DENV vaccination (i.e., >60% seropositivity among <9 year olds), unlike several settings in Latin America and Asia(45). However, nationally representative sampling is necessary before fully applying such criteria in this setting.

Cross-reactivity is a fundamental challenge of ELISA-based serological testing for DENV and CHIKV. Misclassification can arise from cross-reactivity to antibodies derived from infection by, or vaccination against, other flaviviruses for DENV (e.g., yellow fever virus [YFV], West Nile virus, Zika virus, others in circulation) or alphaviruses for CHIKV (e.g., O’nyong’nyong virus [ONNV]). Seroprevalences here represent probable infections, though sensitivity analyses quantifying impact of misclassification generally support our findings. Distinguishing DENV from other flaviviruses remains challenging across tropical regions given frequent co-circulation. In the DRC, YFV is endemic and vaccination is routine in childhood. Zika and West Nile are presumed uncommon in Kinshasa, but limited surveillance precludes exclusion of their contribution(19,22). Other understudied flaviviruses may also contribute. Use of three immunoassays increases confidence that DENV is common in this cohort and a relevant public health problem, while also indicating likely non-DENV flavivirus exposure. For CHIKV, ONNV seroprevalence in Kinshasa is uncertain; no human infections are documented to-date, though virus has been detected among mosquito pools(19).

This study has several limitations. FOI models are based on cross-sectional data, do not include transmission-related covariates, and assume no sero-reversion or heterotypic DENV infection, potentially underestimating transmission. As such, FOI models serve as a first but informative step in characterizing DENV and CHIKV infection risks in this population. Accordingly, FOI reconstructions reflect modeled summaries of historical transmission rather than direct measures of temporal changes in incidence. We also assume infections were acquired at study sites, potentially over-or-under estimating site-specific heterogeneity if infections occurred elsewhere. Low self-reported travel (22%) makes this less likely. Lastly, seroprevalences are not directly generalizable to the entire DRC or surrounding region. Nonetheless, they are an important step toward filling knowledge gaps in arboviral impact within this understudied region.

Our findings reveal a history of sustained, underappreciated DENV transmission in Kinshasa Province and highlight substantial DENV and CHIKV IgG seropositivity, with distinct transmission patterns and spatial heterogeneity. Results underscore need for strengthened arboviral surveillance and diagnostic capacity in the DRC. Improved understanding of arboviral epidemiology is needed to guide management of non-malarial febrile illness, *Aedes-*targeted vector control, and consideration of DENV and CHIKV vaccines and other interventions as they become available.

## ACKNOWLEDGEMENTS

We thank the study team for conducting household surveys and field sampling, as well as all study participants for their time and engagement throughout multiple years of follow-up. We also acknowledge key contributions to design of the parent studies by Steven R. Meshnick (deceased). The authors used artificial intelligence language models for coding support and manuscript text editing. However, the manuscript is original to the authors, who take responsibility for its content.

## BIOGRAPHICAL SKETCH

Rachel Sendor is a PhD candidate in epidemiology at the University of North Carolina at Chapel Hill. Her primary research interests include infectious disease and pharmacoepidemiology, with a focus on vector-borne diseases.

## DATA AVAILABILITY

Data will be made available through the Carolina Digital Repository.

