## Supplemental Materials for "Dengue and chikungunya virus transmission in Kinshasa, Democratic Republic of the Congo"

**Table of Contents**

| | |
|---|---|
| S Figure 1 | Participant flow diagram |
| S Text | Data collection details |
| S Table 2 | Specified force of infection models |
| S Table 3 | Missing data summary by health area |
| S Table 4 | List of R packages used for analysis |
| S Figure 2 | Flow of participants into the validation population, and validation DENV IgG testing |
| S Table 5 | Comparison of DENV serological testing between whole virus ELISA and E-dimer ELISA, among validation sub-population (n=260) |
| S Table 6 | DENV IgG seroprevalence among samples “doubly-ELISA seropositive” in validation population |
| S Figure 3 | Correlates of DENV IgG seroprevalence, stratified by health area |
| S Figure 4 | Correlates of CHIKV IgG seroprevalence, stratified by health area |
| S Table 7 | Sensitivity analysis results – DENV IgG seroprevalence by varying false positivity percent |
| S Table 8 | Sensitivity analysis results – DENV Force of infection by varying false positivity percent |
| S Table 9 | Sensitivity analysis results – CHIKV IgG seroprevalence by varying false positivity percent |
| S Table 10 | Sensitivity analysis results – CHIKV Force of infection by varying false positivity percent |
| S Figure 5 | DENV and CHIKV constant transmission, FOI and age-stratified seroprevalences |
| S Figure 6 | Impact of Kimpoko on increasing DENV FOI in time-varying models |

**S Figure 1: Participant Flow Diagram**

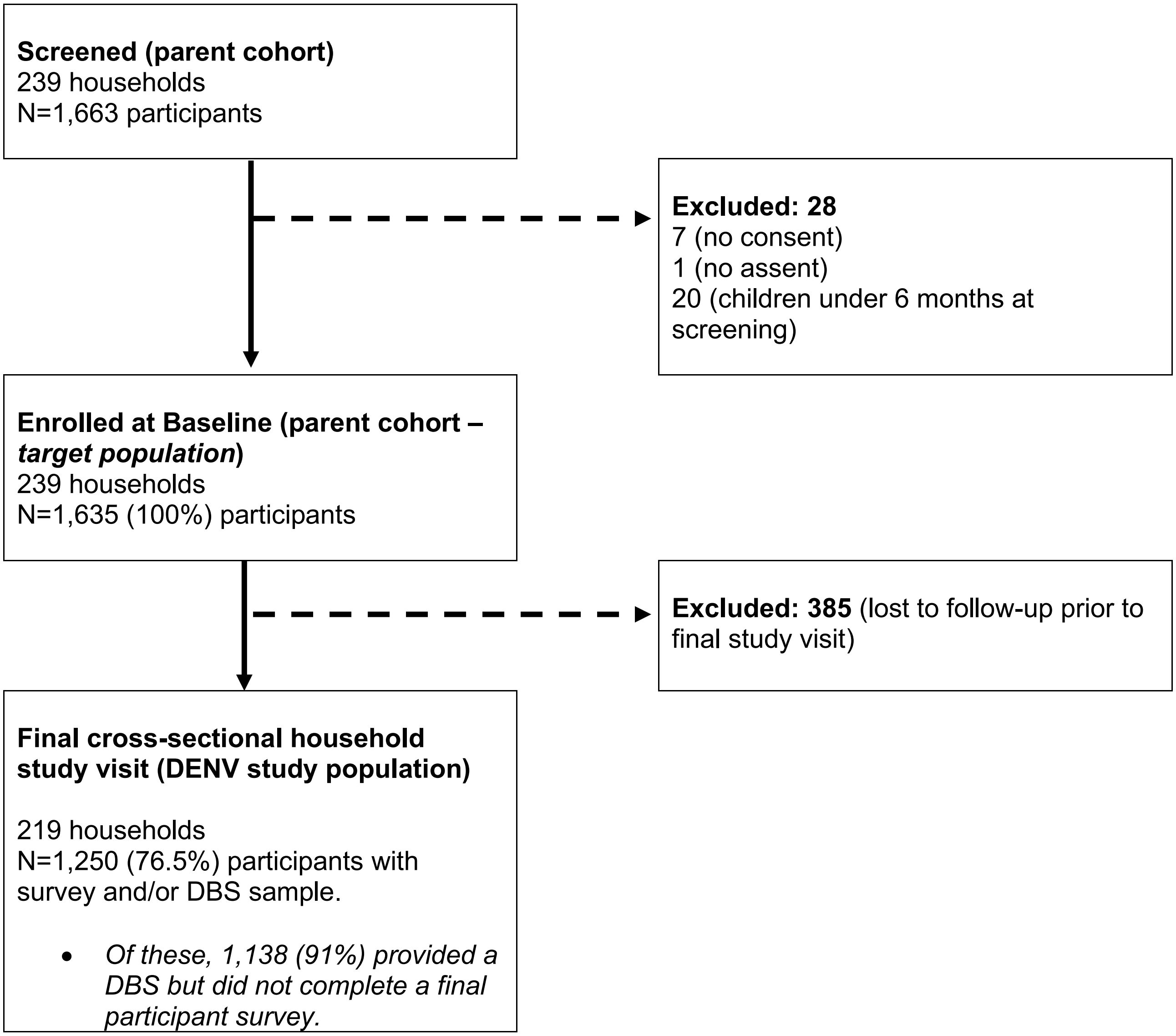

Participant enrollment and retention within the Phase II Kinshasa Malaria Cohort study that gave rise to the analytic sample population for this study is described in detail in Kashamuka et al. *Baseline characteristics for phase II of the Kinshasa Malaria Cohort study: cohort profile*. *BMJ Open, 2024:*14: e085360.

**Supplemental Text: Additional Data Collection Details**

| |
|---|
| Participant survey and DBS data collection<br><br>Participants were surveyed during interviews by a local field team member in their local preferred language. Interviewer-administered questionnaires captured information on demographics, household characteristics and possessions, travel history, and participant behavioral and clinical characteristics. Household-level data was only collected at baseline and was assumed to be constant for the full study period (e.g., housing materials, household possessions, land ownership and water sources, as well as education level and occupation). Dried blood spots (DBS) used in the present study analysis were collected at time of sampling using Whatman® 3MM filter paper (Fisher Scientific, Fair Lawn, NJ USA) and were stored with desiccant at -80°C for future testing. |
| Wealth variable derivation<br><br>Wealth was calculated by household, using household-level possession and housing quality indicators from the baseline survey within a principal component analysis (PCA), generally following the method used in the WHO Demographics and Health Surveys (DHS). Wealth scores were estimated among the entire population, then grouped into quintiles ranging from poorest to wealthiest household; because wealth quintiles were computed in the total population, and not within each health area or village, health area and village are correlated with wealth quintile. |
| Travel history variable derivation<br><br>Recent travel history (defined on the survey as self-reported travel outside of the participant's resident village or Kinshasa within the past 2 weeks prior to the visit) was added partway through the parent cohort baseline, at follow-up visits 4, 5, and 6 (final visit). It was defined as a covariate in our analysis using responses from all available study visits, in order to gain an understanding of historical travel behavior. We therefore classified recent travel history as any reported history of travel outside the village within the follow-up visits where the response was captured, not just the cross-sectional visit giving rise to the analytic sample population. |

*Additional details on data collection methods from the parent cohort giving rise to the study analysis cross-sectional population.*

**S Table 2: Specified FOI modeling equations**

| Model type | Model equation | Assumptions |
|---|---|---|
| **Constant transmission *(across calendar time and age strata)*** | $$P_{(t)} = 1 - (1 - P_0)e^{-\lambda t}$$<br>Where:<br>- *t,* represents calendar time since birth, and<br>- $P_0$ represents the seroprevalence at time, *t=0*, (i.e. birth), which is assumed to be 0. | All individuals are susceptible at birth (i.e. no maternal antibodies).<br><br>Constancy in transmission risk over calendar time within yearly intervals.<br><br>No sero-reversion (i.e. lifelong antibodies with no waning immunity)<br><br>Log-normal distribution (mean = 0.01, sd = 1) prior assumed for DENV and CHIKV FOI, and restricted to values >0.<br><br>Binomial likelihood specified. |
| **Time-varying transmission *(across calendar time)*** | $$P_{(a,t)} = 1 - (1 - P_0)e^{-(\sum_k \lambda_k \Delta t_k)}$$<br>Where:<br>- *a*, represents the age of the individual at, *T*, time of survey (i.e. 2021),<br>- *k,* represents the 5-year time intervals from time, *T*, to time, *T-a,*<br>- $\Delta t_k$, represents the person-years the individual contributed within each, *k*, interval,<br>- $P_0$ represents the seroprevalence at time, *t=0*, (i.e. birth), which is assumed to be 0.<br><br>Summing the product of the force of infection, λ, and person-years within each interval, $\Delta t_k$, across all *k* intervals estimates the cumulative hazard. | All individuals are susceptible at birth (i.e. no maternal antibodies).<br><br>Transmission risk allowed to vary by calendar time, though is assumed 'piece-wise constant' and smoothed across 5-year intervals from 1930 (*corresponding to the oldest age participant*) through 2021 (*the year of the serological survey*).<br><br>Transmission is assumed to be constant within defined age strata.<br><br>No sero-reversion (i.e. lifelong antibodies with no waning immunity<br><br>Log-normal distribution (mean = 0.01, sd = 1) prior assumed for DENV and CHIKV FOI, and restricted to values >0;<br><br>Priors were adjusted according to total population FOI for stratified models due to lower sample size in select age strata.<br><br>Binomial likelihood specified.<br><br>Random-walk smoothing parameters were implemented to correct divergent chains and support construction of earlier time periods. |

Source: *Serofoi* R package vignette documentation: https://github.com/epiverse-trace/serofoi/tree/main/vignettes

**S Table 3: Missing Data Summary, overall and stratified by health area**

| Participant Characteristics | Total Study Population | Voix du Peuple (Urban) | Kimpoko (Peri-urban) | Bu (Rural) |
|---|---|---|---|---|
| **N (%)** | 1,250 (100) | 287 (23.0) | 488 (39.0) | 475 (38.0) |
| **Urbanicity** | | | | |
| Rural | 475 (38.0) | 0 | 0 | 475 (38.0) |
| Peri-urban | 488 (39.0) | 0 | 488 (39.0) | 0 |
| Urban | 287 (23.0) | 287 (23.0) | 0 | 0 |
| Missing | 0 | 0 | 0 | 0 |
| **Village site** | | | | |
| Bu | 163 (13.0) | 0 | 0 | 163 (13.0) |
| Impuru | 166 (13.3) | 0 | 0 | 166 (13.3) |
| Pema | 146 (11.7) | 0 | 0 | 146 (11.7) |
| Ngamanzo | 201 (16.1) | 0 | 201 (16.1) | 0 |
| Iye | 107 (8.6) | 0 | 107 (8.6) | 0 |
| Kimpoko | 180 (14.4) | 0 | 180 (14.4) | 0 |
| Voix du peuple | 287 (23.0) | 287 (23.0) | | 0 |
| Missing | 0 | 0 | 0 | 0 |
| **Sex** | | | | |
| Female | 676 (54.1) | 164 (57.1) | 248 (50.8) | 264 (55.6) |
| Male | 574 (45.9) | 123 (42.9) | 240 (49.2) | 211 (44.4) |
| Missing | 0 | 0 | 0 | 0 |
| **Age (yrs)** | | | | |
| Median [IQR] | 18 [10, 38] | 21 [15, 36] | 17.5 [10, 39] | 14 [8, 37] |
| 3-9 | 290 (23.2) | 35 (12.2) | 102 (20.9) | 153 (32.2) |
| 10-25 | 508 (40.6) | 132 (46.0) | 216 (44.3) | 160 (33.7) |
| 26-50 | 290 (23.2) | 82 (28.6) | 104 (21.3) | 104 (21.9) |
| 51+ | 162 (13.0) | 38 (13.2) | 66 (13.5) | 58 (12.2) |
| Missing | 0 | 0 | 0 | 0 |
| **SES (wealth) score** | | | | |
| Poorest | 237 (19.0) | 0 | 76 (15.6) | 161 (33.9) |
| Poorer | 240 (19.2) | 0 | 82 (16.8) | 158 (33.3) |
| Average wealth | 256 (20.5) | 0 | 136 (27.9) | 120 (25.3) |
| Wealthier | 271 (21.7) | 47 (16.4) | 188 (38.5) | 36 (7.6) |
| Wealthiest | 246 (19.7) | 240 (83.6) | 6 (1.2) | 0 |
| Missing | 0 | 0 | 0 | 0 |
| **Num. household members** | | | | |
| Median [IQR] | 7 [5, 9] | 8 [6, 9] | 6 [5, 8] | 7 [5, 9] |
| 0-5 | 404 (32.3) | 60 (20.9) | 182 (37.3) | 162 (34.1) |
| 6-10 | 691 (55.3) | 173 (60.3) | 257 (52.7) | 261 (54.9) |
| 11+ | 155 (12.4) | 54 (18.8) | 49 (10.0) | 52 (10.9) |
| Missing | 0 | 0 | 0 | 0 |
| **Mosquito nets in household** | | | | |
| Yes | 981 (78.5) | 220 (76.7) | 393 (80.5) | 368 (77.5) |
| No | 269 (21.5) | 67 (23.3) | 95 (19.5) | 107 (22.5) |
| Missing | 0 | 0 | 0 | 0 |

**S Table 3: Missing Data Summary, overall and stratified by health area**

| Participant Characteristics | Total Study Population | Voix du Peuple (Urban) | Kimpoko (Peri-urban) | Bu (Rural) |
|---|---|---|---|---|
| **Num. mosquito nets in household** | | | | |
| Median [IQR] | 2 [1, 3] | 2 [1, 3] | 2 [1, 3] | 2 [1, 3] |
| 0 | 269 (21.5) | 67 (23.3) | 95 (19.5) | 107 (22.5) |
| 1-3 | 835 (66.8) | 175 (61.0) | 325 (66.6) | 335 (70.5) |
| 4+ | 146 (11.7) | 45 (15.7) | 68 (13.9) | 33 (6.9) |
| Missing | 0 | 0 | 0 | 0 |
| **Water w/in 2 min. walk** | | | | |
| Yes | 243 (19.4) | 72 (25.1) | 59 (12.1) | 112 (23.6) |
| No | 1007 (80.6) | 215 (74.9) | 429 (87.9) | 363 (76.4) |
| Missing | 0 | 0 | 0 | 0 |
| **Type of water source** | | | | |
| Stream | 54 (4.3) | 3 (1.0) | 43 (8.8) | 8 (1.7) |
| Pond/lake | 26 (2.1) | 0 | 0 | 26 (5.5) |
| Swamp | 13 (1.0) | 0 | 0 | 13 (2.7) |
| Frequent puddles | 96 (7.7) | 23 (8.0) | 8 (1.6) | 65 (13.7) |
| Other | 30 (2.4) | 22 (7.7) | 8 (1.6) | 0 |
| No water source | 1007 (80.6) | 215 (74.9) | 429 (87.9) | 363 (76.4) |
| Missing | 24 (1.9) | 24 (8.4) | 0 | 0 |
| **Open eaves/holes in home** | | | | |
| Yes | 347 (27.8) | 20 (7.0) | 137 (28.1) | 190 (40.0) |
| No | 894 (71.5) | 267 (93.0) | 342 (70.1) | 285 (60.0) |
| Missing | 9 (0.7) | 0 | 9 (1.8) | 0 |
| **Owns cultivable land** | | | | |
| Yes | 345 (27.6) | 4 (1.4) | 89 (18.2) | 252 (53.1) |
| No | 905 (72.4) | 283 (98.6) | 399 (81.8) | 223 (46.9) |
| Missing | 0 | 0 | 0 | 0 |
| **House has windows** | | | | |
| Yes | 1179 (94.3) | 287 (100) | 459 (94.1) | 433 (91.2) |
| No | 3 (0.2) | 0 | 0 | 3 (0.6) |
| Missing | 68 (5.4) | 0 | 29 (5.9) | 39 (8.2) |
| **Window material** | | | | |
| Glass | 196 (15.7) | 169 (58.9) | 23 (4.7) | 4 (0.8) |
| Screens | 9 (0.7) | 0 | 0 | 9 (1.9) |
| Plastic | 38 (3.0) | 0 | 0 | 38 (8.0) |
| Planks | 480 (38.4) | 59 (20.6) | 225 (46.1) | 196 (41.3) |
| Open holes | 152 (12.2) | 3 (1.0) | 42 (8.6) | 107 (22.5) |
| Other | 264 (21.1) | 57 (19.9) | 142 (29.1) | 65 (13.7) |
| No windows | 3 (0.2) | 0 | 0 | 3 (0.6) |
| Missing | 46 (3.7) | 0 | 28 | 18 |
| **Education Attained** | | | | |
| No school | 89 (7.1) | 7 (2.4) | 31 (6.4) | 51 (10.7) |
| Primary | 406 (32.5) | 58 (20.2) | 158 (32.4) | 190 (40.0) |
| Secondary / Post-secondary | 499 (39.9) | 187 (65.2) | 196 (40.2) | 116 (24.4) |

| S Table 3: Missing Data Summary, overall and stratified by health area | | | | |
|---|---|---|---|---|
| **Participant Characteristics** | **Total Study Population** | **Voix du Peuple (Urban)** | **Kimpoko (Peri-urban)** | **Bu (Rural)** |
| Other | 27 (2.2) | 6 (2.1) | 11 (2.3) | 10 (2.1) |
| Under school-age (<5 yrs) | 59 (4.7) | 6 (2.1) | 19 (3.9) | 34 (7.2) |
| Missing | 170 (13.6) | 23 (8.0) | 73 (15.0) | 74 (15.6) |
| **History of travel** | | | | |
| Yes | 272 (21.8) | 25 (8.7) | 106 (21.7) | 141 (29.7) |
| No | 954 (76.3) | 250 (87.1) | 375 (76.8) | 329 (69.3) |
| Missing | 24 (1.9) | 12 (4.2) | 7 (1.4) | 5 (1.1) |
| **CHIKV IgG** | | | | |
| Yes | 324 (26.6) | 14 (5.1) | 154 (32.0) | 156 (32.7) |
| No | 916 (73.3) | 269 (93.7) | 329 (67.4) | 318 (66.9) |
| Missing | 10 (0.8) | 4 (1.4) | 5 (1.0) | 1 (0.2) |
| **DENV_IgG** | | | | |
| Yes | 502 (40.2) | 92 (32.1) | 282 (57.8) | 128 (26.9) |
| No | 738 (59.0) | 191 (66.6) | 201 (41.2) | 346 (72.8) |
| Missing | 10 (0.8) | 4 (1.4) | 5 (1.0) | 1 (0.2) |

| **Covariates with missing data** | **Proportion missing** | **Imputed with MICE?** |
|---|---|---|
| education level | 14% | Y |
| recent travel | 9% | Y |
| house has windows | 5% | Y |
| window material | 4% | N[1] |
| history of any travel | 2% | Y |
| type of water source nearby | 2% | Y |
| DENV IgG | 0.8% | Y |
| CHIKV IgG | 0.8% | Y |
| eaves in home | 0.7% | Y |

[1] Window material was not imputed as it was not a mutually exclusive field.

**S Table 4: List of R packages used for analysis**

| R package | Version | Citation |
|---|---|---|
| broom | 1.0.3 | Robinson D, Hayes A, Couch S (2023). *broom: Convert Statistical Objects into Tidy Tibbles*. R package version 1.0.3, https://CRAN.R-project.org/package=broom. |
| car | 3.1.1 | Fox J, Weisberg S (2019). *An R Companion to Applied Regression*, Third edition. Sage, Thousand Oaks CA. https://socialsciences.mcmaster.ca/jfox/Books/Companion/. |
| devtools | 2.4.5 | Wickham H, Hester J, Chang W, Bryan J (2022). *devtools: Tools to Make Developing R Packages Easier*. R package version 2.4.5, https://CRAN.R-project.org/package=devtools. |
| dplyr | 1.1.4 | Wickham H, François R, Henry L, Müller K, Vaughan D (2023). *dplyr: A Grammar of Data Manipulation*. R package version 1.1.4, https://CRAN.R-project.org/package=dplyr. |
| forestplot | 3.1.7 | Gordon M, Lumley T (2025). *forestplot: Advanced Forest Plot Using 'grid' Graphics*. R package version 3.1.7, https://CRAN.R-project.org/package=forestplot. |
| gee | 4.13.25 | Carey VJ (2022). *gee: Generalized Estimation Equation Solver*. R package version 4.13-25, https://CRAN.R-project.org/package=gee. |
| geepack | 1.3.9 | Halekoh U, Højsgaard S, Yan J (2006). “The R Package geepack for Generalized Estimating Equations.” *Journal of Statistical Software*, 15(2), 1-11. Yan J, Fine JP (2004). “Estimating Equations for Association Structures.” *Statistics in Medicine*, 23, 859-880. Yan J (2002). “geepack: Yet Another Package for Generalized Estimating Equations.” *R-News*, 2(3), 12-14. |
| ggExtra | 0.10.0 | Attali D, Baker C (2022). *ggExtra: Add Marginal Histograms to 'ggplot2', and More 'ggplot2' Enhancements*. R package version 0.10.0, https://CRAN.R-project.org/package=ggExtra. |
| ggplot2 | 3.5.2 | Wickham H (2016). *ggplot2: Elegant Graphics for Data Analysis*. Springer-Verlag New York. ISBN 978-3-319-24277-4, https://ggplot2.tidyverse.org. |
| janitor | 2.2.0 | Firke S (2023). *janitor: Simple Tools for Examining and Cleaning Dirty Data*. R package version 2.2.0, https://CRAN.R-project.org/package=janitor. |
| kableExtra | 1.4.0 | Zhu H (2024). *kableExtra: Construct Complex Table with 'kable' and Pipe Syntax*. R package version 1.4.0, https://CRAN.R-project.org/package=kableExtra. |
| linelist | 0.0.1 | Jombart T (2022). *linelist: Tagging and Validating Epidemiological Data*. R package version 0.0.1, https://CRAN.R-project.org/package=linelist. |
| lmtest | 0.9.40 | Zeileis A, Hothorn T (2002). “Diagnostic Checking in Regression Relationships.” *R News*, 2(3), 7-10. https://CRAN.R-project.org/doc/Rnews/. |
| leaflet | 2.2.3. | Cheng J, Schloerke B, Karambelkar B, Xie Y, Aden-Buie G (2025).leaflet: Create Interactive Web Maps with the JavaScript 'Leaflet' R package version 2.2.3, <https://CRAN.R-project.org/package=leaflet>. |
| mice | 3.16.0 | van Buuren S, Groothuis-Oudshoorn K (2011). “mice: Multivariate Imputation by Chained Equations in R.” *Journal of Statistical Software*, 45(3), 1-67. doi:10.18637/jss.v045.i03 https://doi.org/10.18637/jss.v045.i03. |
| miceadds | 3.17.44 | Robitzsch A, Grund S (2024). *miceadds: Some Additional Multiple Imputation Functions, Especially for 'mice'*. R package version 3.17-44, https://CRAN.R-project.org/package=miceadds. |
| mitools | 2.4 | Lumley T (2019). *mitools: Tools for Multiple Imputation of Missing Data*. R package version 2.4, https://CRAN.R-project.org/package=mitools. |
| plyr | 1.8.9 | Wickham H (2011). “The Split-Apply-Combine Strategy for Data Analysis.” *Journal of Statistical Software*, 40(1), 1-29. https://www.jstatsoft.org/v40/i01/. |
| PropCIs | 0.3.0 | Scherer R (2018). *PropCIs: Various Confidence Interval Methods for Proportions*. R package version 0.3-0, https://CRAN.R-project.org/package=PropCIs. |
| purrr | 1.0.2 | Wickham H, Henry L (2023). *purrr: Functional Programming Tools*. R package version 1.0.2, https://CRAN.R-project.org/package=purrr. |

**S Table 4: List of R packages used for analysis**

| R package | Version | Citation |
|---|---|---|
| readr | 2.1.3 | Wickham H, Hester J, Bryan J (2022). *readr: Read Rectangular Text Data*. R package version 2.1.3, https://CRAN.R-project.org/package=readr. |
| readxl | 1.4.1 | Wickham H, Bryan J (2022). *readxl: Read Excel Files*. R package version 1.4.1, https://CRAN.R-project.org/package=readxl. |
| rstan | 2.32.6 | Stan Development Team (2024). *RStan: the R interface to Stan*. R package version 2.32.6, https://mc-stan.org/. |
| sandwich | 3.1.1 | Zeileis A, Köll S, Graham N (2020). “Various Versatile Variances: An Object-Oriented Implementation of Clustered Covariances in R.” *Journal of Statistical Software*, 95(1), 1-36. doi:10.18637/jss.v095.i01 https://doi.org/10.18637/jss.v095.i01. Zeileis A (2004). “Econometric Computing with HC and HAC Covariance Matrix Estimators.” *Journal of Statistical Software*, 11(10), 1-17. doi:10.18637/jss.v011.i10 https://doi.org/10.18637/jss.v011.i10. Zeileis A (2006). “Object-Oriented Computation of Sandwich Estimators.” *Journal of Statistical Software*, 16(9), 1-16. doi:10.18637/jss.v016.i09 https://doi.org/10.18637/jss.v016.i09. |
| serofoi | 1.0.3 | Cucunubá Z, Domínguez NT, Lambert B, Nouvellet P (2025). *serofoi: Bayesian Estimation of the Force of Infection from Serological Data*. https://github.com/epiverse-trace/serofoi, https://epiverse-trace.github.io/serofoi/. |
| sf | 1.0.10 | Pebesma E (2018). “Simple Features for R: Standardized Support for Spatial Vector Data.” *The R Journal*, 10(1), 439-446. doi:10.32614/RJ-2018-009 https://doi.org/10.32614/RJ-2018-009. Pebesma E, Bivand R (2023). *Spatial Data Science: With applications in R*. Chapman and Hall/CRC. https://r-spatial.org/book/. |
| splines | 4.2.2 | R Core Team (2022). *R: A Language and Environment for Statistical Computing*. R Foundation for Statistical Computing, Vienna, Austria. https://www.R-project.org/. |
| stats | 4.2.2 | R Core Team (2022). *R: A Language and Environment for Statistical Computing*. R Foundation for Statistical Computing, Vienna, Austria. https://www.R-project.org/. |
| stringr | 1.5.1 | Wickham H (2023). *stringr: Simple, Consistent Wrappers for Common String Operations*. R package version 1.5.1, https://CRAN.R-project.org/package=stringr. |
| survey | 4.4.2 | Lumley T (2024). *survey: analysis of complex survey samples*. R package version 4.4. Lumley T (2004). “Analysis of Complex Survey Samples.” *Journal of Statistical Software*, 9(1), 1-19. Lumley T (2010). *Complex Surveys: A Guide to Analysis Using R*. John Wiley and Sons. |
| tableone | 0.13.2 | Yoshida K, Bartel A (2022). *tableone: Create 'Table 1' to Describe Baseline Characteristics with or without Propensity Score Weights*. R package version 0.13.2, https://CRAN.R-project.org/package=tableone. |
| tidyr | 1.3.1 | Wickham H, Vaughan D, Girlich M (2024). *tidyr: Tidy Messy Data*. R package version 1.3.1, https://CRAN.R-project.org/package=tidyr. |
| tidyverse | 1.3.2 | Wickham H, Averick M, Bryan J, Chang W, McGowan LD, François R, Grolemund G, Hayes A, Henry L, Hester J, Kuhn M, Pedersen TL, Miller E, Bache SM, Müller K, Ooms J, Robinson D, Seidel DP, Spinu V, Takahashi K, Vaughan D, Wilke C, Woo K, Yutani H (2019). “Welcome to the tidyverse.” *Journal of Open Source Software*, 4(43), 1686. doi:10.21105/joss.01686 https://doi.org/10.21105/joss.01686. |
| VIM | 6.2.2 | Kowarik A, Templ M (2016). “Imputation with the R Package VIM.” *Journal of Statistical Software*, 74(7), 1-16. doi:10.18637/jss.v074.i07 https://doi.org/10.18637/jss.v074.i07. |
| knitr | 1.5 | Xie Y (2025). *knitr: A General-Purpose Package for Dynamic Report Generation in R*. R package version 1.50, https://yihui.org/knitr/. Xie Y (2015). *Dynamic Documents with R and knitr*, 2nd edition. Chapman and Hall/CRC, Boca Raton, Florida. ISBN 978-1498716963. Xie Y (2014). “knitr: A Comprehensive Tool for Reproducible Research in R.” In Stodden V, Leisch F, Peng RD (eds.), *Implementing Reproducible Computational Research*. Chapman and Hall/CRC. ISBN 978-1466561595. |

**S Figure 2. Flow of participants into the Validation Population, and validation DENV IgG testing**

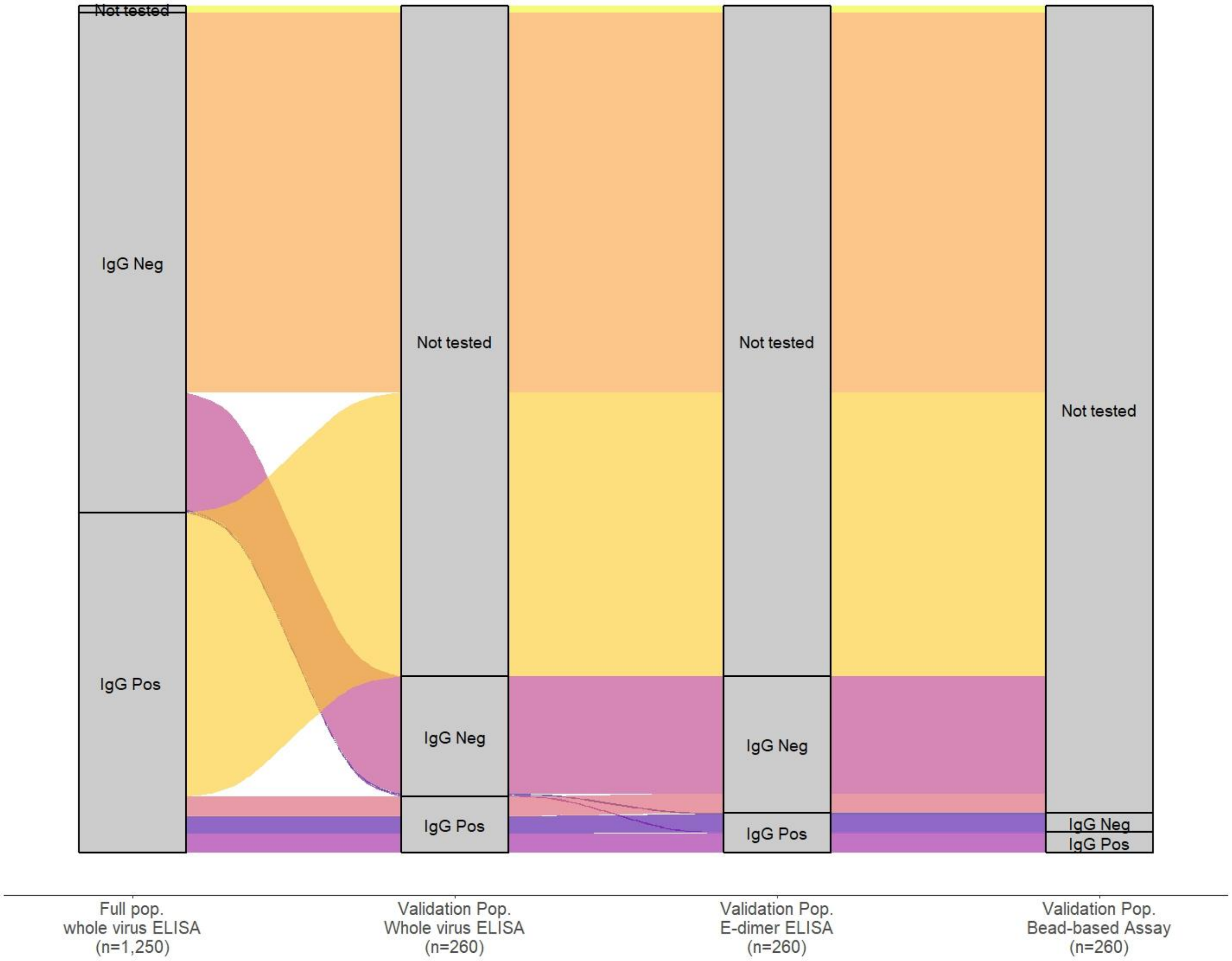

| | Full Population | Validation Population (n=260) | | |
|---|---|---|---|---|
| | Whole virus ELISA | Whole virus ELISA | E-dimer ELISA | Bead-based Assay |
| | N (%) | N (% [of 260 tested]) | N (% [of 260 tested]) | N (% of 58 tested[1]) |
| IgG Pos | 479 (38.3%) | 83 (32%) | 59 (23%) | 30 (51.7%)[3] |
| IgG Neg | 761 (60.9%) | 177 (68%) | 201 (77%) | 28 (48.3%) |
| Not tested | 10 (0.8%) | 990 | 990 | 990 + 202[2] = 1,192 |
| Total | 1,250 | 1,250 | 1,250 | 1,250 |

[1] 58 of the 59 E-dimer ELISA IgG seropositives were further tested by bead-based assay for DENV confirmation. N=1 E-dimer ELISA seropositive sample was not tested.

[2] E-dimer ELISA IgG seronegatives + 1 untested E-dimer seropositive sample.

[3] 28 of these 30 IgG seropositive samples by BBA were IgG+ across all three assays – whole virus ELISA, E-dimer ELISA, and BBA.

**S Table 5. Comparison of DENV serological testing between whole virus ELISA and E-dimer ELISA, among validation sub-population (n=260)**

| | | Whole virus ELISA | | |
|---|---|---|---|---|
| | | **DENV IgG +** | **DENV IgG -** | **TOTAL** |
| **E-dimer ELISA** | **DENV IgG +** | 55 | 4 | 59 |
| | **DENV IgG -** | 28 | 173 | 201 |
| | **TOTAL** | 83 | 177 | 260 |
| **Concordance** | | | | |
| Overall | | | (173+55) / 260 | **87.7%** |
| DENV IgG + | | | 55 / 83 | **66.3%** |
| DENV IgG - | | | 173/177 | **97.7%** |
| **Cohen's Kappa** | | | | **0.693** |

**S Table 6. DENV IgG seroprevalence among samples "doubly-ELISA seropositive" in validation population**

| | **Denv IgG+ 'doubly-seropositives'** *IgG+ by whole virus ELISA and E-dimer ELISA* | | | |
|---|---|---|---|---|
| | **N events[1]** | **N total** | **Age-standardized seroprevalence[2]** | **95% CI** |
| **Total** | 55 | 260 | 30.2% | 22.7% - 37.6% |
| **Age (years)** | | | | |
| 3-6 | 4 | 72 | 5.5% | 0.23% - 10.9% |
| 7-10 | 18 | 98 | 18.4% | 10.7% - 26.1% |
| 11-25 | 14 | 45 | 31.1% | 17.4% - 44.8% |
| 26-50 | 12 | 29 | 41.4% | 23.1% - 59.6% |
| 51+ | 7 | 16 | 43.8% | 18.6% - 68.9% |
| **Health area** | | | | |
| Voix du Peuple (urban) | 7 | 48 | 19.1% | 6.1% - 32.2% |
| Kimpoko (peri-urban) | 38 | 108 | 37.2% | 26.4% - 47.9% |
| Bu (rural) | 10 | 104 | 8.0% | 2.7% - 13.2% |
| **Village site** | | | | |
| Bu | 4 | 39 | 8.7% | 0.037% - 17.4% |
| Impuru | 5 | 42 | 8.5% | 1.2% - 15.7% |
| Pema | 1 | 23 | 5.0% | -3.1% - 13.1% |
| Ngamanzo | 21 | 38 | 54.5% | 37.4% - 71.6% |
| Iye | 1 | 23 | 4.0% | -0.29% - 10.8% |
| Kimpoko | 16 | 47 | 41.80% | 27.4% - 56.2% |
| Voix du Peuple | 7 | 48 | 19.10% | 6.1% - 32.2% |

[1] samples Denv IgG+ on whole virus antigen-capture ELISA, and also on the recombinant E-dimer ELISA, for higher-confidence dengue calls.

[2] Seroprevalences were age-standardized to account for the age-stratified random sampling scheme used to select the validation sub-population.

**S Figure 3. Correlates of DENV IgG+ Seroprevalence, stratified by health area**

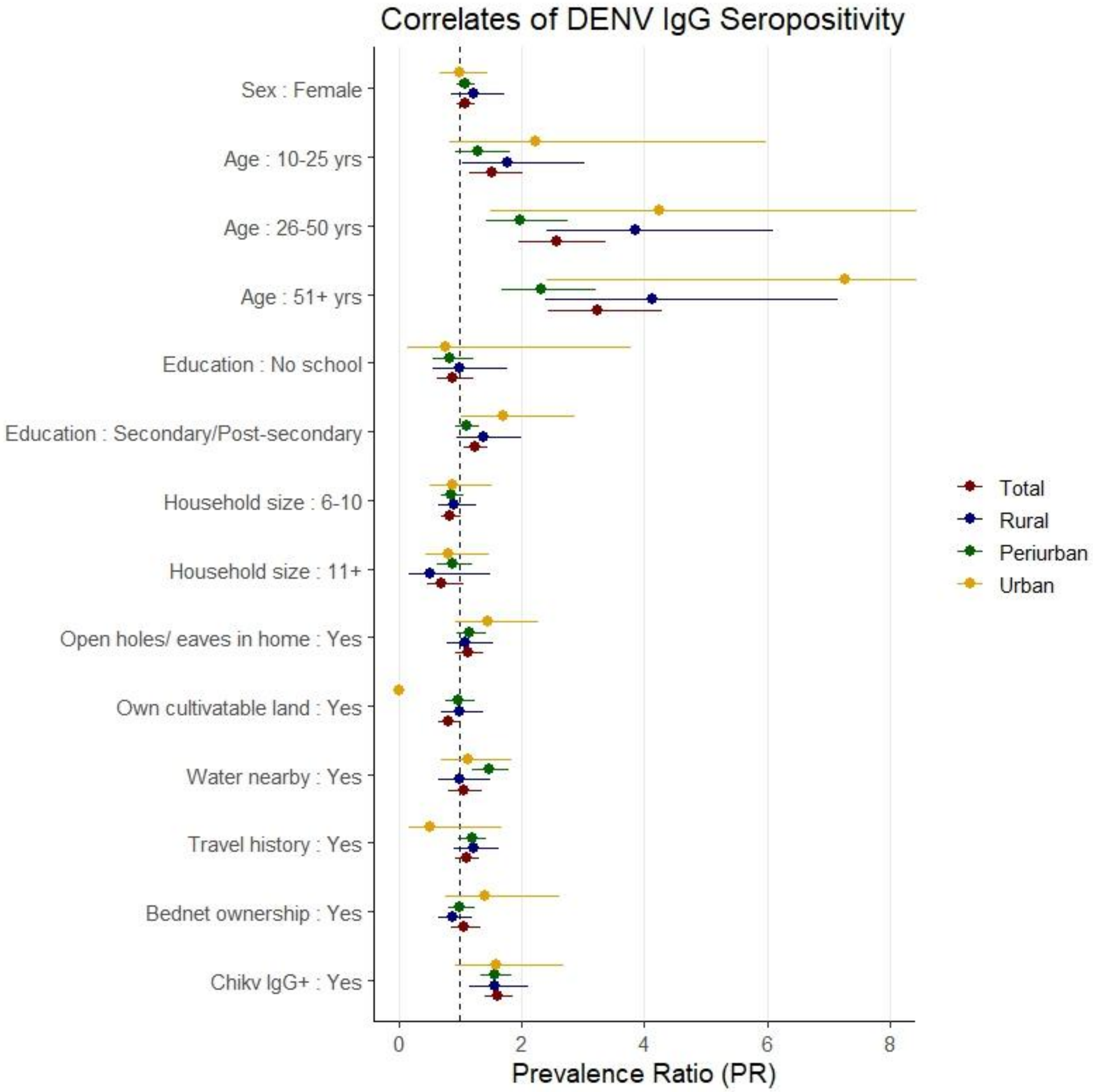

**S Figure 4. Correlates of CHIKV IgG seroprevalence, stratified by health area**

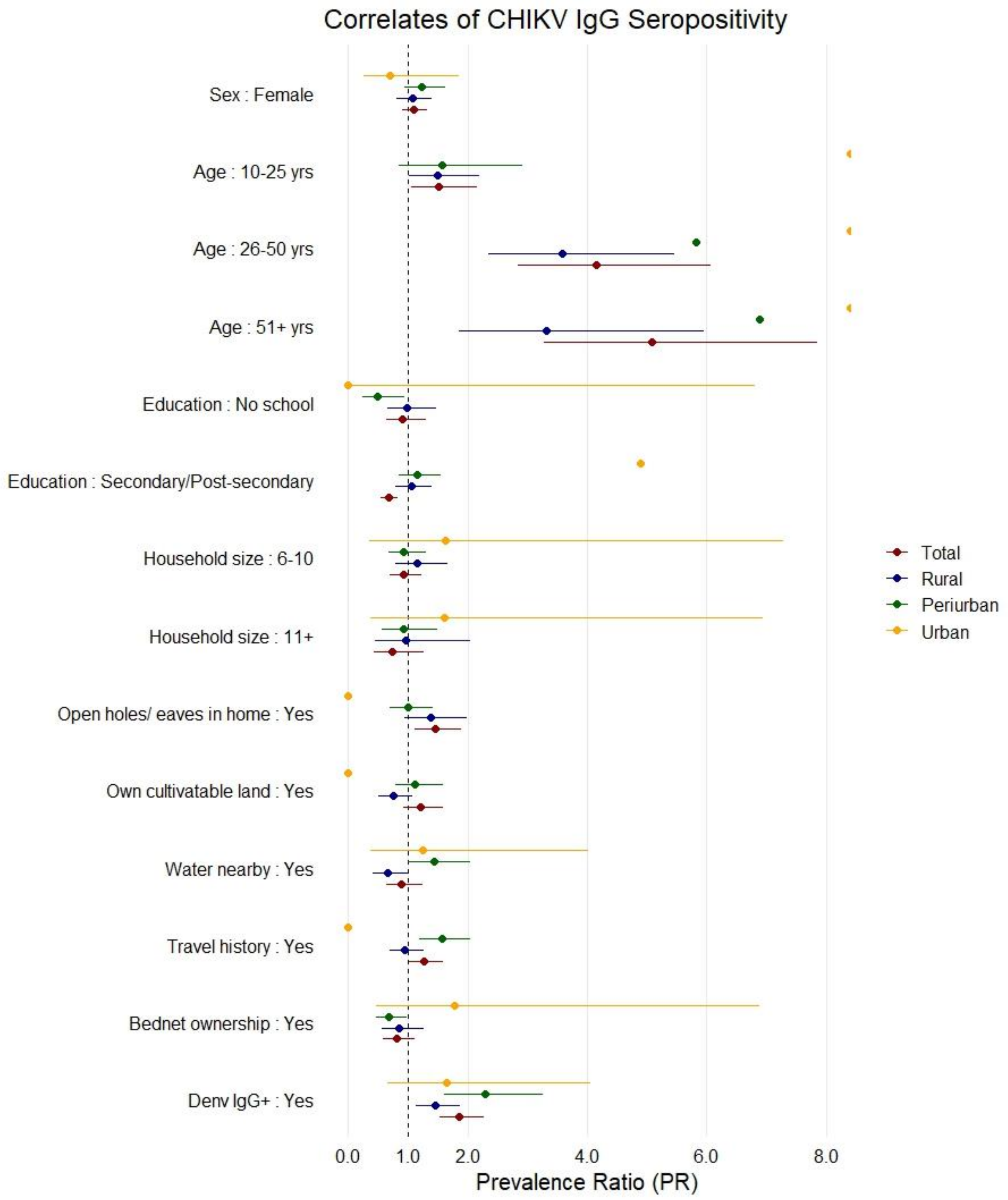

| S Table 7: Sensitivity analysis: DENV IgG seroprevalence by varying false positivity percent | | | | | | | |
|---|---|---|---|---|---|---|---|
| **% False Positivity Assumed** | **Observed IgG+ n** | **Assumed false positives, n** | **DENV IgG+, n** | **DENV IgG-, n** | **TOTAL, N** | **Crude Seroprevalence % (95% CI)** | **Weighted* Seroprevalence (95% CI)** |
| **0% (primary results)** | | | | | | | |
| Total Pop. | 479 | 0 | 479 | 761 | 1,240 | 38.6 % (35.0% - 42.4%) | 37.4% (33.8% - 41.2%) |
| Voix du Peuple (urban) | 88 | 0 | 88 | 195 | 283 | 31.1% (24.8% - 38.2%) | 29.5% (23.3% - 36.6%) |
| Kimpoko (peri-urban) | 271 | 0 | 271 | 212 | 483 | 56.1% (50.7% - 61.3%) | 54.6% (49.1% - 59.9%) |
| Bu (rural) | 120 | 0 | 120 | 354 | 474 | 25.3% (21.3% - 29.7%) | 24.2% (20.4% - 28.6%) |
| **20% false positives** | | | | | | | |
| Total Pop. | 479 | 95 | 384 | 856 | 1,240 | 31.0% (27.8% - 34.3%) | 30.2% (27.0% - 33.5%) |
| Voix du Peuple (urban) | 88 | 19 | 69 | 214 | 283 | 24.4% (18.7% - 31.1%) | 23.3% (17.8% - 29.9%) |
| Kimpoko (peri-urban) | 271 | 49 | 222 | 261 | 483 | 46.0% (41.2% - 50.8%) | 44.8% (40.1% - 49.7%) |
| Bu (rural) | 120 | 27 | 93 | 381 | 474 | 19.6% (16.1% - 23.7%) | 18.9% (15.5% - 22.9%) |
| **30% false positives** | | | | | | | |
| Total Pop. | 479 | 143 | 336 | 904 | 1,240 | 27.1% (24.2% - 30.2%) | 26.5% (23.6% - 29.6%) |
| Voix du Peuple (urban) | 88 | 23 | 65 | 218 | 283 | 23.0% (17.4% - 29.6%) | 22.0% (16.6% - 28.5%) |
| Kimpoko (peri-urban) | 271 | 81 | 190 | 293 | 483 | 39.3% (34.6% - 44.3%) | 38.5% (33.8% - 43.5%) |
| Bu (rural) | 120 | 39 | 81 | 393 | 474 | 17.1% (13.9% - 20.9%) | 16.5% (13.4% - 20.2%) |
| **40% false positives** | | | | | | | |
| Total Pop. | 479 | 191 | 288 | 952 | 1,240 | 23.2% (20.4% - 26.3%) | 22.8% (20.0% - 25.8%) |
| Voix du Peuple (urban) | 88 | 34 | 54 | 229 | 283 | 19.1% (13.9% - 25.6%) | 18.2% (13.2% - 24.5%) |
| Kimpoko (peri-urban) | 271 | 105 | 166 | 317 | 483 | 34.4% (29.7% - 39.3%) | 33.7% (29.1% - 38.7%) |
| Bu (rural) | 120 | 52 | 68 | 406 | 474 | 14.3% (11.3% - 18.0%) | 14.0% (11.0% - 17.5%) |

[1] Non-imputed dataset for sensitivity analysis, with random sampling at designated false positivity percentages to re-classify outcomes at random. Total n=1,240 due to 10 missing dengue virus outcomes.
[2] Weighted to the baseline population to be more representative of the target population in Kinshasa. Household clustering was not accounted for in sensitivity seroprevalence estimation by binomial 1-sample proportion testing.

| S Table 8: Sensitivity analysis results – DENV Force of infection by varying false positivity percent | | |
|---|---|---|
| | **Force of Infection Median (95% CrI)** | |
| | Constant transmission model | Time-varying transmission model |
| **0% (primary results)** | | |
| Total Pop. | 0.022 (0.020 – 0.025) | *1980*: 0.013 (0.005 – 0.027)<br>*2000*: 0.019 (0.010 – 0.034)<br>*2020*: 0.032 (0.023 – 0.044) |
| Voix du Peuple (urban) | 0.015 (0.012 – 0.019) | *1980:* 0.019 (0.008 – 0.037)<br>*2000:* 0.017 (0.009 – 0.028)<br>*2020:* 0.013 (0.006 – 0.020) |
| Kimpoko (peri-urban) | 0.041 (0.036 – 0.046) | *1980:* 0.028 (0.010 – 0.047)<br>*2000:* 0.035 (0.018 – 0.055)<br>*2020:* 0.053 (0.040 – 0.080) |
| Bu (rural) | 0.014 (0.012 – 0.017) | *1980:* 0.009 (0.003 – 0.015)<br>*2000:* 0.013 (0.006 – 0.021)<br>*2020:* 0.017 (0.012 – 0.026) |
| **20% false positives** | | |
| Total Pop. | 0.016 (0.015 – 0.018) | *1980:* 0.005 (0.001 – 0.014)<br>*2000:* 0.012 (0.004 – 0.025)<br>*2020:* 0.027 (0.019 – 0.039) |
| Voix du Peuple (urban) | 0.011 (0.009 – 0.014) | *1980:* 0.004 (0.0004 – 0.016)<br>*2000:* 0.006 (0.001 – 0.018)<br>*2020:* 0.006 (0.001 – 0.015) |
| Kimpoko (peri-urban) | 0.028 (0.024 – 0.031) | *1980:* 0.002 (0.0001 – 0.009)<br>*2000:* 0.006 (0.001 – 0.016)<br>*2020:* 0.031 (0.018 – 0.048) |
| Bu (rural) | 0.010 (0.008 – 0.012) | *1980:* 0.001 (0.000 – 0.005)<br>*2000:* 0.003 (0.0003 – 0.008)<br>*2020:* 0.010 (0.004 – 0.018) |
| **30% false positives** | | |
| Total Pop. | 0.014 (0.012 – 0.015) | *1980:* 0.003 (0.0001 – 0.010)<br>*2000:* 0.008 (0.002 – 0.020)<br>*2020:* 0.026 (0.017 – 0.036) |
| Voix du Peuple (urban) | 0.010 (0.008 – 0.013) | *1980:* 0.005 (0.0005 – 0.016)<br>*2000:* 0.006 (0.001 – 0.017)<br>*2020:* 0.006 (0.001 – 0.015) |
| Kimpoko (peri-urban) | 0.022 (0.019 – 0.025) | *1980:* 0.002 (0.0002 – 0.009)<br>*2000:* 0.006 (0.001 – 0.016)<br>*2020:* 0.030 (0.018 – 0.047) |
| Bu (rural) | 0.009 (0.007 – 0.011) | *1980:* 0.001 (0.000 – 0.005)<br>*2000:* 0.003 (0.0003 – 0.008)<br>*2020:* 0.010 (0.004 – 0.018) |
| **40% false positives** | | |
| Total Pop. | 0.011 (0.010 – 0.013) | *1980:* 0.002 (0.000 – 0.008)<br>*2000:* 0.006 (0.0008 – 0.015)<br>*2020:* 0.023 (0.015 – 0.033) |
| Voix du Peuple (urban) | 0.008 (0.006 – 0.011) | *1980:* 0.005 (0.0006 – 0.016)<br>*2000:* 0.006 (0.002 – 0.017)<br>*2020:* 0.006 (0.001 – 0.015) |
| Kimpoko (peri-urban) | 0.018 (0.015 – 0.021) | *1980:* 0.002 (0.0002 – 0.009)<br>*2000:* 0.006 (0.001 – 0.015)<br>*2020:* 0.030 (0.018 – 0.047) |
| Bu (rural) | 0.007 (0.006 – 0.009) | *1980:* 0.001 (0.000 – 0.005)<br>*2000:* 0.003 (0.0004 – 0.008)<br>*2020:* 0.010 (0.004 – 0.018) |

| S Table 9: Sensitivity analysis: CHIKV IgG seroprevalence by varying false positivity percent | | | | | | | |
|---|---|---|---|---|---|---|---|
| **% False Positivity Assumed** | **Observed IgG+ n** | **Assumed false positives, n** | **CHIKV IgG+, n** | **CHIKV IgG-, n** | **TOTAL, N** | **Crude Seroprevalence % (95% CI)** | **Weighted* Seroprevalence % (95% CI)** |
| **0% (primary results)** | | | | | | | |
| Total Pop. | 324 | 0 | 324 | 916 | 1,240 | 26.1% (22.8% - 29.7%) | 24.4% (21.2% - 28.0%) |
| Voix du Peuple (urban) | 14 | 0 | 14 | 269 | 283 | 5.0% (3.0% - 8.1%) | 4.6% (2.8% - 7.5%) |
| Kimpoko (peri-urban) | 154 | 0 | 154 | 329 | 483 | 31.9% (27.3% - 36.8%) | 29.9% (25.5% - 34.8%) |
| Bu (rural) | 156 | 0 | 156 | 318 | 474 | 32.9% (27.4% - 29.0%) | 31.8% (26.2% - 38.1%) |
| **20% false positives** | | | | | | | |
| Total Pop. | 324 | 64 | 260 | 980 | 1,240 | 21.0% (18.2% - 24.0%) | 19.6% (16.9% - 22.5%) |
| Voix du Peuple (urban) | 14 | 4 | 10 | 273 | 283 | 3.5% (2.0% - 6.0%) | 3.3% (1.9% - 5.7%) |
| Kimpoko (peri-urban) | 154 | 27 | 127 | 356 | 483 | 26.3% (22.3% - 30.7%) | 24.6% (20.8% - 28.9%) |
| Bu (rural) | 156 | 33 | 123 | 351 | 474 | 25.9% (21.4% - 31.1%) | 25.0% (20.4% - 20.1%) |
| **30% false positives** | | | | | | | |
| Total Pop. | 324 | 97 | 227 | 1013 | 1,240 | 18.3% (15.8% - 21.1%) | 17.1% (14.7% - 19.9%) |
| Voix du Peuple (urban) | 14 | 4 | 10 | 273 | 283 | 3.5% (2.0% - 6.0%) | 3.3% (1.9% - 5.7%) |
| Kimpoko (peri-urban) | 154 | 43 | 111 | 372 | 483 | 23.0% (19.1% - 27.4%) | 21.7% (18.0% - 25.9%) |
| Bu (rural) | 156 | 50 | 106 | 368 | 474 | 22.4% (18.4% - 26.9%) | 21.5% (17.6% - 26.1%) |
| **40% false positives** | | | | | | | |
| Total Pop. | 324 | 129 | 195 | 1045 | 1,240 | 15.7% (13.5% - 18.2%) | 14.7% (12.6% - 17.2%) |
| Voix du Peuple (urban) | 14 | 6 | 8 | 275 | 283 | 2.8% (1.5% - 5.2%) | 2.7% (1.5% - 5.0%) |
| Kimpoko (peri-urban) | 154 | 59 | 95 | 388 | 483 | 19.7% (16.2% - 23.6%) | 18.6% (15.3% - 22.4%) |
| Bu (rural) | 156 | 64 | 92 | 382 | 474 | 19.4% (15.8% - 23.6%) | 18.6% (15.1% - 22.8%) |

[1] Non-imputed dataset for sensitivity analysis, with random sampling at designated false positivity percentages to re-classify outcomes at random. Total n=1,240 due to 10 missing dengue virus outcomes.
[2] Weighted to the baseline population to be more representative of the target population in Kinshasa. Household clustering was not accounted for in sensitivity seroprevalence estimation by binomial 1-sample proportion testing.

| S Table 10: Sensitivity analysis results – CHIKV Force of infection by varying false positivity percent | | |
|---|---|---|
| | **Force of Infection Median (95% Crl)** | |
| | Constant transmission model | Time-varying transmission model |
| **0% (primary results)** | | |
| Total Pop. | 0.013<br>(0.012 – 0.015) | *1980*: 0.012 (0.0005 – 0.044)<br>*2000*: 0.007 (0.0004 – 0.023)<br>*2020*: 0.026 (0.016 – 0.035) |
| Voix du Peuple (urban) | 0.002<br>(0.001 – 0.003) | *1980*: 0.002 (0.0005 – 0.006)<br>*2000*: 0.002 (0.0005 – 0.004)<br>*2020*: 0.001 (0.0003 – 0.004) |
| Kimpoko (peri-urban) | 0.018<br>(0.015 – 0.021) | *1980*: 0.025 (0.008 – 0.055)<br>*2000*: 0.022 (0.010 – 0.044)<br>*2020*: 0.015 (0.009 – 0.024) |
| Bu (rural) | 0.020<br>(0.017 – 0.023) | *1980*: 0.012 (0.004 – 0.028)<br>*2000*: 0.015 (0.006 – 0.029)<br>*2020*: 0.029 (0.020 – 0.044) |
| **20% false positives** | | |
| Total Pop. | 0.010<br>(0.009 – 0.012) | *1980*: 0.007 (0.0002 – 0.026)<br>*2000*: 0.007 (0.0003 – 0.020)<br>*2020*: 0.018 (0.011 – 0.026) |
| Voix du Peuple (urban) | 0.001<br>(0.0007 – 0.002) | *1980*: 0.001 (0.0003 – 0.004)<br>*2000*: 0.001 (0.0003 – 0.003)<br>*2020*: 0.001 (0.0003 – 0.003) |
| Kimpoko (peri-urban) | 0.014<br>(0.011 – 0.016) | *1980*: 0.016 (0.005 – 0.032)<br>*2000*: 0.017 (0.009 – 0.039)<br>*2020*: 0.011 (0.006 – 0.017) |
| Bu (rural) | 0.015<br>(0.012 – 0.017) | *1980*: 0.011 (0.003 – 0.025)<br>*2000*: 0.012 (0.004 – 0.021)<br>*2020*: 0.020 (0.013 – 0.032) |
| **30% false positives** | | |
| Total Pop. | 0.009<br>(0.008 – 0.010) | *1980*: 0.007 (0.0003 – 0.019)<br>*2000*: 0.006 (0.0005 – 0.016)<br>*2020*: 0.015 (0.009 – 0.023) |
| Voix du Peuple (urban) | 0.001<br>(0.001 – 0.002) | *1980*: 0.002 (0.0003 – 0.004)<br>*2000*: 0.001 (0.0004 – 0.003)<br>*2020*: 0.001 (0.0003 – 0.003) |
| Kimpoko (peri-urban) | 0.011<br>(0.010 – 0.014) | *1980*: 0.011 (0.003 – 0.022)<br>*2000*: 0.013 (0.007 – 0.030)<br>*2020*: 0.010 (0.005 – 0.017) |
| Bu (rural) | 0.012<br>(0.010 – 0.015) | *1980*: 0.009 (0.003 – 0.021)<br>*2000*: 0.009 (0.003 – 0.017)<br>*2020*: 0.017 (0.011 – 0.027) |
| **40% false positives** | | |
| Total Pop. | 0.007<br>(0.006 – 0.008) | *1980*: 0.006 (0.0005 – 0.015)<br>*2000*: 0.006 (0.0008 – 0.013)<br>*2020*: 0.011 (0.007 – 0.019) |
| Voix du Peuple (urban) | 0.001<br>(0.001 – 0.002) | *1980*: 0.001 (0.0003 – 0.003)<br>*2000*: 0.001 (0.0003 – 0.003)<br>*2020*: 0.001 (0.0003 – 0.003) |
| Kimpoko (peri-urban) | 0.010<br>(0.008 – 0.012) | *1980*: 0.009 (0.003 – 0.016)<br>*2000*: 0.010 (0.006 – 0.022)<br>*2020*: 0.009 (0.005 – 0.014) |
| Bu (rural) | 0.010<br>(0.008 – 0.012) | *1980*: 0.008 (0.003 – 0.017)<br>*2000*: 0.008 (0.003 – 0.013)<br>*2020*: 0.013 (0.009 – 0.022) |

**S Figure 5: Dengue virus (A-D) and chikungunya virus (E-H) Age-stratified seroprevalence curves and FOI models, assuming constant transmission over time and age. ).** Seroprevalence within 10-year age strata is plotted on the primary y-axis for observed (*points and 95% CIs*) and posterior-predicted (*solid line and 95% CrIs*) values for DENV (A-D) and CHIKV (E-H). Force of infection (FOI) predicted from posterior distributions (*dashed line and 95% CrIs*) is plotted on the secondary y-axis for: total population (**red**); downtown Kinshasa site (**yellow**); Kimpoko health area sites (**light green**); Bu health area sites (**blue**). Constant transmission models (**a-d**) apply a catalytic modeling function and assume constant transmission risk across calendar time with no waning immunity among infected individuals.

***Dengue virus (A-D)***

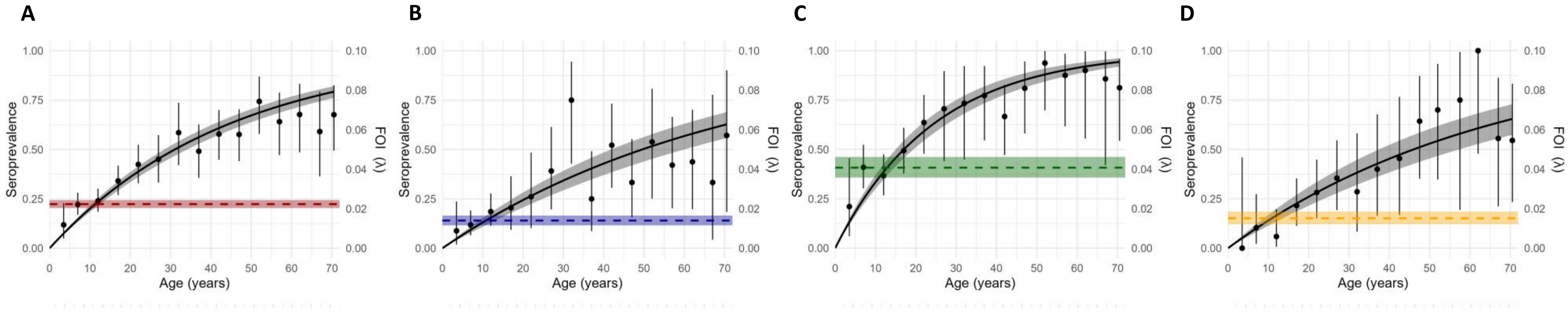

***Chikungunya virus (E-H)***

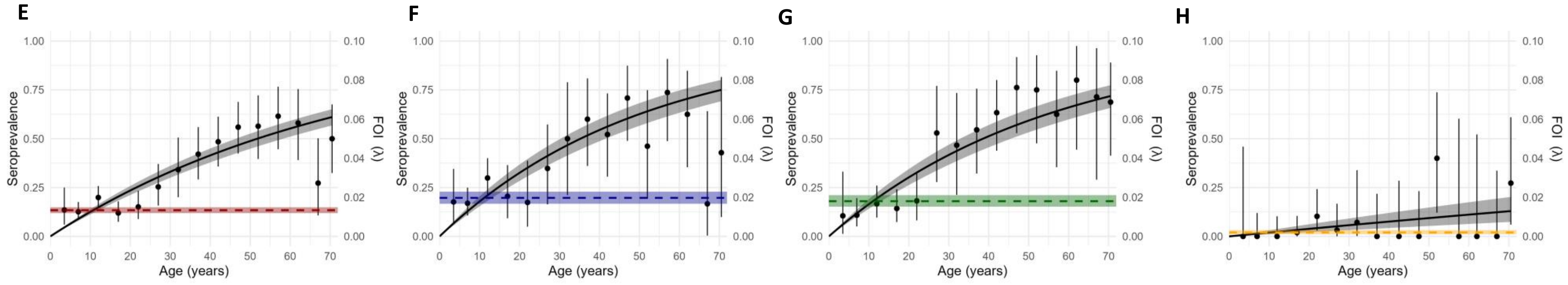

**S Figure 6**

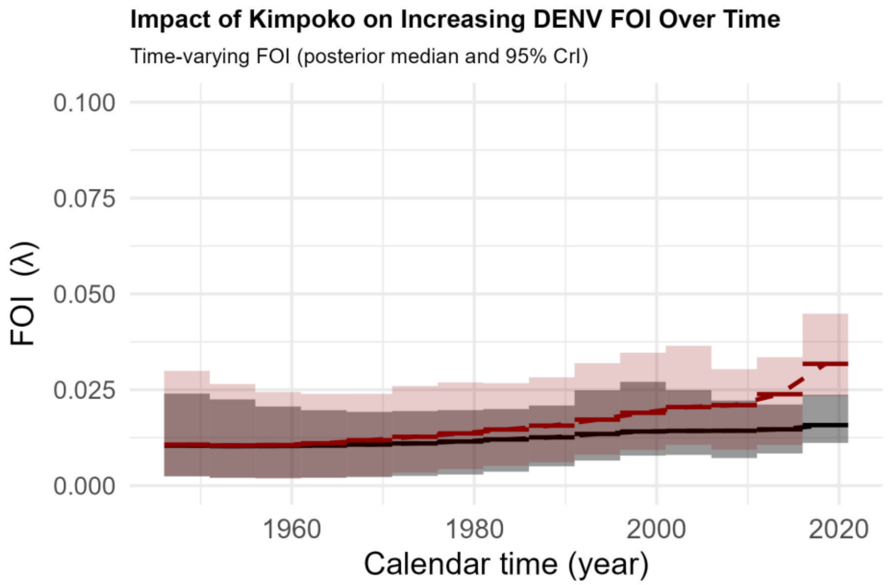

**Red line:** Total population, including Kimpoko health area; **Black line:** Population excluding Kimpoko health area.
